# Findings and insights from the genetic investigation of age of first reported occurrence for complex disorders in the UK Biobank and FinnGen

**DOI:** 10.1101/2020.11.20.20234302

**Authors:** Yen-Chen A. Feng, Tian Ge, Mattia Cordioli, FinnGen, Andrea Ganna, Jordan W. Smoller, Benjamin M. Neale

**Author notes:** Correspondence: Jordan W. Smoller, Benjamin M. Neale.

## Abstract

Age of onset contains information on the timing of events relevant to disease etiology, but there has not been a systematic investigation of its heritability from GWAS data. Here, we characterize the genetic architecture of age of first occurrence and its genomic relationship with disease susceptibility for a wide range of complex disorders in the UK Biobank. For diseases with a sufficient sample size, we discover that age of first occurrence has non-trivial genetic contributions, some with specific genetic risk factors not associated with susceptibility to the disease. Through genetic correlation analysis, we show that an earlier health-event occurrence is correlated with a higher polygenic risk of disease susceptibility. An independent genetic investigation of the FinnGen cohort replicates the pattern of heritability and genetic correlation estimates. We then demonstrate that incorporating disease onset age with susceptibility may improve genetic risk prediction and stratification.

## Introduction

Genome-wide association studies (GWAS) have revealed that most complex traits and diseases have an underlying polygenic component^1,2^. To date, disease-based GWAS have predominantly examined the genetic risk associated with *whether* an individual has ever been affected with a disease of interest (i.e., disease susceptibility) using a case-control design. Such design, however, typically uses lifetime risk to model the association of phenotypic variation with genetic variation and ignores the time component of *when* a disease occurs for an individual.

Studies have shown that age of onset of a disease itself plays a pertinent role in understanding the genetic etiology of disease development. Using a genome-wide approach, efforts have been made to identify genetic modifiers of disease onset age^3-5^, as well as genetic risk factors distinctive to different age-of-onset groups^6-8^. As shown from twin and single-nucleotide-polymorphism (SNP) heritability analyses, the phenotypic variance explained by genetic variation can change across the lifespan for many traits^9-11^. Furthermore, the polygenic model has suggested that there may exist a correlation between age of onset and susceptibility to a disease, such that individuals with a higher genetic loading might develop the disease at an earlier age^12-14^. Despite these efforts, however, there has been comparatively less investigation of the heritability of age of onset for disease phenotypes.

In recent years, large-scale biobank datasets that include broad phenotypic information (e.g., UK Biobank^15,16^, FinnGen^17,18^) have provided an unprecedented resource to study the causes of disease at scale with a linkage to genetic data. Related to age of onset, dates of health-outcome events, such as diagnosis, treatment, and death, have been made available through questionnaire-based or hospital records within such biobanks. Collectively, this provides an opportunity to systematically characterize the genetic construct of disease onset age, the specific genetic risk factors that may alter it, and how these findings correlate or differ from the genetic basis of susceptibility.

Here, we present a deep investigation into the genetic architecture of age of first occurrence and its genetic overlap with susceptibility across a wide range of complex disorders in the UK Biobank (UKBB). We leveraged measurements from three different phenotypic datasets that either directly or approximately capture age of onset: *self-reported* age of diagnosis of a medical condition (SR), age of first in-patient ICD-10 diagnosis or hospitalization episode from *hospital in-patient* records (HIP), and age of the earliest event occurrence *combining* self-report, in-patient, primary care, and death records (COMB; Methods). We refer to these definitions more generally as “age of first occurrence” of a particular disease or medical condition. For diseases with a sufficient sample size, we show that age of first occurrence is moderately heritable, some with specific genetic risk factors not associated with susceptibility. Across disease domains, there is an overall inverse genetic correlation between age of first occurrence and susceptibility. Independent of the UKBB cohort, we then show that a similar pattern of heritability and genetic correlations exists in the FinnGen study, which has a longer follow-up. Finally, we demonstrate that information on age of first occurrence has the potential to improve polygenic risk prediction for disease susceptibility and patient stratification.

## Results

### Characterizing age of first occurrence in the UK Biobank

We estimated age of first occurrence for medical conditions aggregated from the SR, HIP, and COMB datasets in the UKBB White-British subset (N=361,140). We extracted every possible disease definition following hierarchical disease classifications where appropriate, resulting in over 3,000 clinical terms (Methods). Among them, 70 SR, 224 HIP, and 164 COMB terms had at least 5000 affected individuals (prevalence ∼1.4%; Tables S1-3) and were retained for GWAS analysis.

Age of first occurrence ranged from 0 to 70 years of age in the SR dataset (median: 17-59), 30 to 80 in the HIP dataset (median: 36-66), and 0 to 80 in the COMB dataset (median: 6-68; Figures 1&S1; Tables S4-6). We then compared the distribution of age of first occurrence by data source for 26 disease phenotypes where definitions were comparable between SR and HIP, of which 15 mapped across all three datasets (Table S7). Among these diseases, SR age of first occurrence was consistently younger than that based on the HIP dataset. Some diseases had a higher prevalence in SR (e.g., hypertension, asthma, and arthrosis) while others (e.g., hernia, gallstones, and cancers) were more prevalent in the HIP dataset. Distribution of age of first occurrence in SR varied extensively by trait and exhibited a spiky behavior by quartiles (0.25, 0.5, 0.75, or 1.00), reflecting that age of the reported diagnosis was recorded as integers in the questionnaire. Comparatively, age of first occurrence in HIP was estimated directly from the recorded dates and was smoothly and consistently distributed across diseases with a bell-like shape. The COMB dataset had the largest number of cases per definition among all three sources and showed an overall “merged” distribution of age of first occurrence in SR and HIP (Figures 1B&S1).

**Figure 1.**
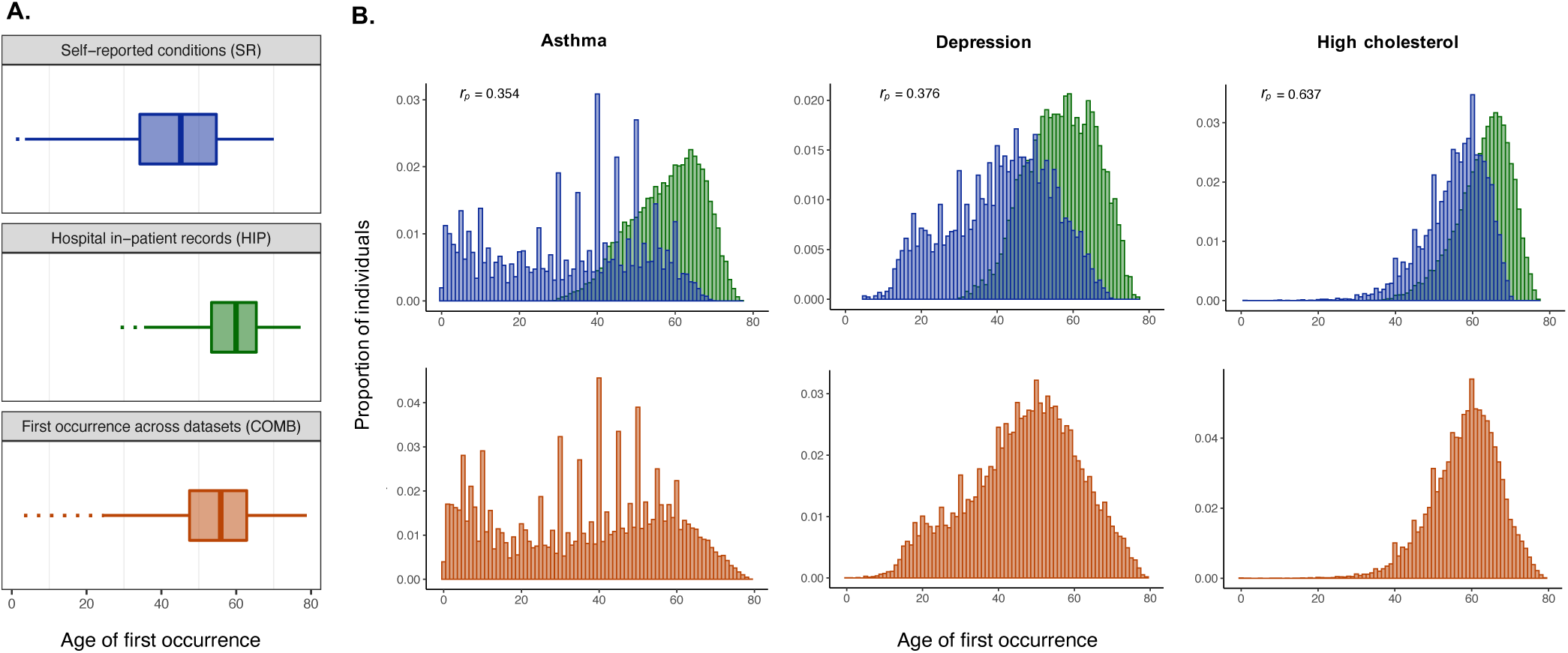
Distribution of age of first occurrence of disease phenotypes from three phenotypic datasets in UKBB. **A**. An averaged distribution of age of first occurrence is shown across 70 SR (blue), 224 HIP (green), and 164 COMB (orange) disease definitions in each dataset. Dotted line indicates the outlying range of values. Age of first occurrence ranges from 0-70 in the SR dataset, 30-80 in the HIP dataset, and 0-80 in the COMB dataset. Spikes in the SR phenotypes reflect that the values are recorded in quartiles (0.25, 0.5, 0.75, or 1.00). **B**. Distribution of age of first occurrence differs by trait and data source. Shown here are three selected disease phenotypes with matching definitions across datasets. SR and HIP conditions show little to moderate overlap in age of first occurrence, as measured by Pearson’s correlation coefficient (*r*_p_; top), while the COMB conditions exhibit a merged distribution of SR and HIP (bottom).

Considering diagnosis of the same medical condition from both SR and HIP, the distribution of age of first occurrence between the two datasets showed little (e.g, migraine, median difference *m*_*diff*_ = 31.6, phenotypic correlation *r*_*p*_ = 0.12; asthma, *m*_*diff*_ = 27.2, *r*_*p*_ = 0.35) to moderate overlap (e.g, high cholesterol, *m*_*diff*_ = 6.4, *r*_*p*_ = 0.64; gallstones, *m*_*diff*_ = 8.6, *r*_*p*_ = 0,87), with values in HIP shifting toward the right (*m*_*diff*_: 6-32; *r*_*p*_: 0.1-0.9; avg. *r*_*p*_ = 0.46; Figures 1B&S2; Table S7). The extent of overlap generally increased with age of first occurrence of the condition (Figure S3) but overall suggested that the actual disease “onset” age for most diseases was left-truncated in the hospitalization records, reflecting the fact that ICD-10 was implemented and integrated in the UK hospital episode statistics in the 1990s. As expected, age of first occurrences in the COMB dataset overlapped substantially with both the SR (avg. *r*_*p*_ = 0.91) and the HIP (avg. *r*_*p*_ = 0.69) datasets (Table S7).

### Common genetic associations with age of first occurrence

We performed a GWAS of age of first occurrence for the 70 SR, 224 HIP, and 164 COMB disease definitions with >5000 affected individuals. For each condition, a case-control GWAS was also performed, treating non-diseased individuals as the control group and adjusting for the same sets of covariates (Methods). Univariate Linkage Disequilibrium Score Regression (LDSR) intercepts for all GWASes had a value close to 1, suggesting little or no signs of inflation in association statistics due to population stratification or other confounding factors (Figure S4; Tables S8-10).

Genome-wide significant SNPs (*P* < 5×10^−8^) that may alter age of first occurrence were identified for 31 SR, 57 HIP, and 42 COMB disease definitions, ranging from 1 to 20 independently associated loci. Among the three data sources, age of first occurrence defined for hospitalization events in the HIP dataset had the least number of associated loci (max. 2 loci; Tables S8-10). While most of the identified signals were a subset of the significant associations in the corresponding susceptibility GWAS (Figures 2A&S5-7), some of these GWASes contained loci significantly associated with age of first occurrence but not with susceptibility, suggesting a role in modifying disease development that precedes disease onset (Tables S8-11).

**Figure 2.**
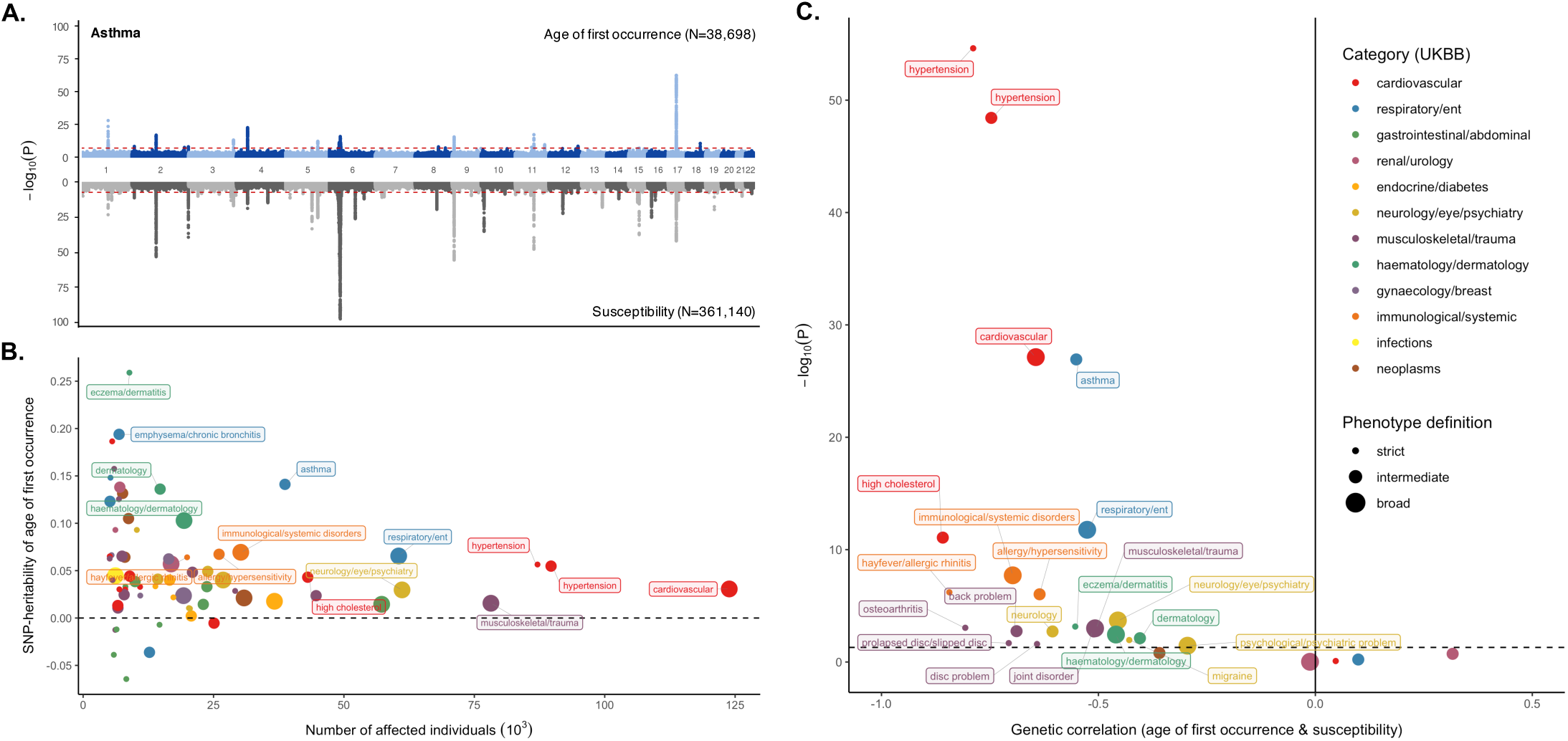
Genetic characterization of age of first occurrence and its relationship with susceptibility in the UKBB SR dataset. **A**. A Miami plot of GWAS results reveal overlapping and distinct genetic associations between age of first occurrence (top) and case-control status (bottom) of SR asthma. Each dot represents a single SNP. P-values are shown on the −log_10_ scale on the y-axis, plotted against chromosome positions on the x-axis. The red dashed lines denote the genome-wide significance threshold at P = 5×10-8. **B**. SNP-heritability estimates for age of first occurrence 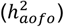across 70 SR disease definitions suggest non-trivial common genetic contributions. 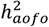 was estimated from univariate LDSR. Each dot represents an individual disease, colored by disease categories used in the SR dataset; a larger dot corresponds to a broader disease definition. Labeled are conditions with a significant 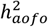 at FDR < 0.05. Heritability analysis of HIP and COMB diseases reveal a similar pattern in Figure S9. **C**. Genetic correlation (rg) analysis suggests an inverse genomic relationship between age of first occurrence and susceptibility for diseases with a significant heritability for both traits. rg was estimated using bivariate LDSR. The dashed line denotes nominal significance at P = 0.05; labeled are conditions with a significant rg at FDR < 0.05. Analysis of HIP and COMB diseases show a similar pattern in Figure S13.

Disease phenotypes showing the largest number of independent associations with age of first occurrence included asthma (total number of significant hits, *n*_*sig*_ = 19; total number of unique hits not seen in susceptibility GWAS, *n*_*uniq*_ = 5), hypertension (*n*_*sig*_ = 9), high cholesterol level (*n*_*sig*_ = 6, *n*_*uniq*_ = 1), and eczema (*n*_*sig*_ =5, *n*_*uniq*_ = 2) in the SR dataset. Top results in the HIP dataset included mental disorders (*n*_*sig*_ = 2, *n*_*uniq*_ = 2), monoarthritis (*n*_*sig*_ = 2 *n*_*uniq*_ = 2), substance-related disorders (*n*_*sig*_ = 2, *n*_*uniq*_ = 2), and type 2 diabetes (*n*_*sig*_ = 2, *n*_*uniq*_ = 1); in the COMB dataset those included asthma (*n*_*sig*_ = 20, *n*_*uniq*_ = 7), disorders of lipoprotein metabolism (*n*_*sig*_ = 13), measles (*n*_*sig*_ = 6, *n*_*uniq*_ = 6), and dermatitis (*n*_*sig*_ = 5, *n*_*uniq*_ = 1) (Tables S8-10).

Comparing the overlapping loci between age of first occurrence and susceptibility revealed that their effects were often in opposite directions and although in close proximity, the lead SNPs were mostly different. A few loci showed an even stronger association with age of first occurrence than with susceptibility. For instance, the significant locus on chromosome 1 for SR asthma was associated with a reduced age of onset by 4.8 years (lead SNP 1:152285861:G:A, *P* = 4.6×10^−29^) but an increased risk of asthma by 1.2 fold (lead SNP 1:152179152:C:T, *P* = 7.4×10^−24^; Table S11).

### Moderate SNP-heritability of age of first occurrence

Heritability analysis using LDSR showed that age of first occurrence for complex diseases was moderately heritable: 27/70 SR endpoints, 49/224 HIP endpoints, and 30/164 COMB endpoints had a significantly non-zero SNP-heritability for the age at which an individual first developed a given condition 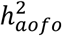; nominal p-value < 0.05), ranging from 1 to 25% with an average of 7-9% (Figures 2B&S8-9; Tables S12-14). 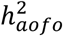 estimates in the HIP dataset were slightly lower than in the other two datasets.

Diseases with a heritable age of first occurrence in the SR dataset included cardiovascular (e.g., hypertension, 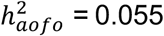, respiratory (e.g., asthma, 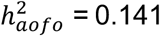, dermatological (e.g., eczema/dermatitis, 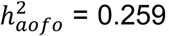 and immunological (e.g., allergy/hypersensitivity/anaphylaxis, 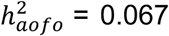 related traits (Figure 2B; see p-values in Table S12). Top diseases in the HIP dataset were seen among circulatory system (e.g., hypertension, 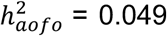, genitourinary (e.g., irregular menstrual cycle/bleeding, 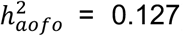, digestive (e.g., cholelithiasis and cholecystitis, 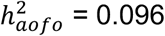, mental disorders (e.g., psychological disorders, 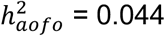, and neoplasms (e.g., benign or malignant skin cancer, 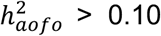 Figure S9A; Table S13). Significant 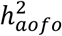 in the COMB dataset encompassed some of the top non-cancer conditions from SR and HIP, as well as other traits like myocardial infarction 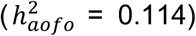non-insulin-dependent diabetes 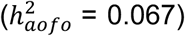, hypothyroidism 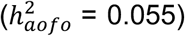and diaphragmatic hernia 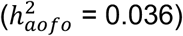 (Figure S9B; Table S14).

Among the 26 mapped disease definitions, 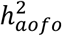 estimates showed variability across datasets, particularly between SR and HIP, with some diseases more consistently estimated than others (e.g., hypertension, 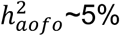; Table S7; Figure S10A). Conditions with a significant 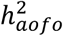 in SR but not in HIP—often also larger in magnitude—tended to be chronic that could start early in life or are mild in presentations (e.g., asthma, disc problem, high cholesterol, and dermatological conditions). Conversely, a significant 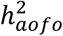 in HIP but not in SR was observed among conditions that appeared more likely to be acute or have an adult-onset that required in-hospital treatments (e.g., gallstones, diabetes, and cancers; Figure S11). Cross-dataset genetic correlation estimates (r_g_) of the mapped age-of-first-occurrence endpoints differed widely and many did not reach statistical significance. On average, these r_g_’s were highest between SR and COMB, lowest between SR and HIP, which increased slightly with age of first occurrence, consistent with the pattern of phenotypic similarity (Table S7; Figure S12 vs. S3).

In contrast, SNP-heritability estimates for the corresponding susceptibility endpoints 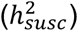 were all significant and showed a lesser degree of heterogeneity across datasets (Figure S10B), with most of the traits having a cross-dataset r_g_ closer to 1 (Tables S7).

## Inverse genetic correlation between age of first occurrence and susceptibility

For diseases with a significant 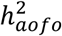 and 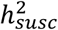, we estimated rg between age of first occurrence and susceptibility using bivariate LDSR (Methods). Interestingly, more than half of the tested traits showed a significant, negative *r*_*g*_ between the risk of developing the disease and the age of developing the disease, ranging from approximately −0.2 to −0.9 (Figures 2C&S13-14; Tables S12-14). This inverse genomic relationship was observed across disease categories in all three phenotypic datasets. A few traits had a non-significant, positive *r*_*g*_ that could result from a smaller sample size (Figure S13).

As with the pattern of 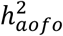, *r*_*g*_ results showed a different profile in each dataset. Hypertension was consistently the most significant phenotype with an *r*_*g*_ for age at first occurrence and susceptibility around −0.7 in all three datasets (*P* < 10^−28^). Other top diseases in the SR dataset with an *r*_*g*_ at FDR < 0.05 included asthma (*r*_*g*_ = −0.55), high cholesterol (*r*_*g*_ = −0.86), hay fever/allergy (*r*_*g*_ < −0.6), eczema (*r*_*g*_ = −0.55), osteoarthritis (*r*_*g*_ = −0.81), and migraine (*r*_*g*_ = −0.43,) (Figure 2C; see *p*-values in Table S12). In the HIP dataset additional traits with the strongest *r*_*g*_ included substance-related/psychological disorders (*r*_*g*_ < −0.3), diseases of esophagus and dysphagia (*r*_*g*_ = −0.79), type 2 diabetes (*r*_*g*_ = −0.51), COPD/bronchiectasis/asthma (*r*_*g*_ = −0.40), non-specific chest pain (*r*_*g*_ = −0.58), and cholelithiasis and cholecystitis (*r*_*g*_ = −0.37) (Figure S14A; Table S13). In the COMB dataset other top results were predominantly a combination of traits from SR and HIP, such as disorders of lipoprotein metabolism (*r*_*g*_ = −0.99), asthma (*r*_*g*_ = −0.56), diverticular disease of intestine (*r*_*g*_ = −0.82), other arthrosis (*r*_*g*_ = −0.72), other hypothyroidism (*r*_*g*_ = −0.72), and non-insulin-dependent diabetes mellitus (*r*_*g*_ = −0.59) (Figure S14B; Table S14). Together, these observations are consistent with predictions of the polygenic model, in which an earlier onset or occurrence of a disease may correlate with a higher polygenic liability^12^.

Across pairs of diseases, *r*_*g*_’s for susceptibility were similar to *r*_*g*_’s for age of first occurrence: that is, diseases with extensive sharing of genetic risk appeared to also have genetically correlated age of first occurrence, especially for phenotypes measured by SR. (Figure S15).

### Similar genetic architecture of age of first occurrence in FinnGen

Next, we sought replication for UKBB findings, specifically the pattern of heritability and genetic correlation, in FinnGen, based on its v4 release of 130,423 unrelated individuals. FinnGen is a registry-based cohort that follows health events across a lifetime for an individual, including medication history (Figure S16; Supplementary Notes). A fraction of the FinnGen participants were ascertained in hospitals or disease-based cohorts^14^, making it a case-enriched cohort relative to UKBB (Table S15). Because registers data were established since the 1960s, FinnGen has the advantage of a prospective cohort that contains a longer follow-up time and a wider age range (0.08 to 98.98 years old at recruitment) for participants compared to UKBB. We analyzed 280 disease phenotypes with a sufficient sample size following the same analytical pipeline in UKBB where possible (Methods; Table S15). Age of first occurrence was defined as the earliest age of an event in the registries (range: 0-100; median: 9-72; Table S16; Figure S17).

As in the UKBB, some diseases were found to have genetic risk factors associated with age of first occurrence but not with susceptibility (e.g., diabetes; Table S17). 63 of the 280 medical conditions had a significant heritability for age of first occurrence, ranging from 2 to 22%, many of which were among top results in UKBB (Figure 3A; Table S18). These included conditions identical to or close proxies for phenotypes in UKBB, particular those defined in the HIP dataset, such as hypertension 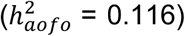, arthropathies 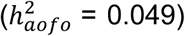, statin medication 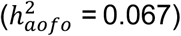 diabetes 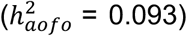, cholelithiasis 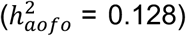, and migraine (triptan purchase and/or ICD diagnosis; 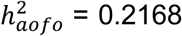. Conditions with a significant 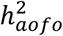 in FinnGen but not in UKBB involved pregnancy and childbirth 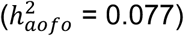 and eye diseases (e.g., senile cataract, 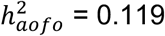 Figure 3A; see p-values in Table S18).

**Figure 3.**
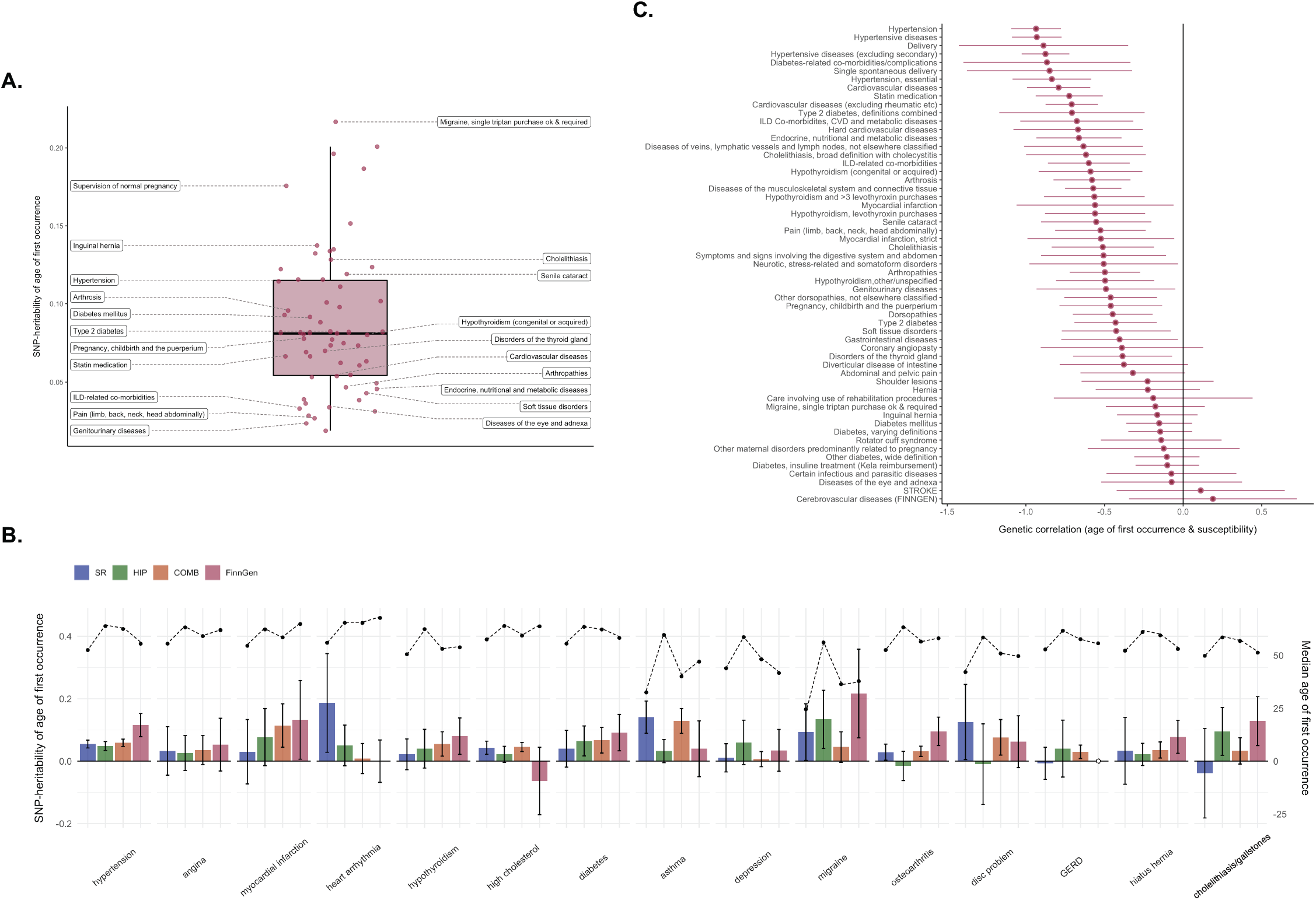
Genetic analysis of age of first occurrence in FinnGen and its comparison with UKBB results. **A**. Distribution of heritability estimates from FinnGen for 64 diseases that have a significant 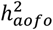. Labeled are selected conditions with a significant 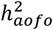 at FDR < 0.05. **B**. 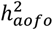 estimates for 15 comparable disease definitions in UKBB and FinnGen show variable degree of similarity. Left axis denotes SNP-heritability shown in bar plots and the corresponding 95% confidence intervals (95% CI). Right axis shows the median age of first occurrence for each condition indicated in dotted lines. The full comparison of all 26 matched phenotypes is available in Table S18 and Figure S19. **C**. A negative *r*_*g*_ between age of first occurrence and disease susceptibility is observed for many of the tested diseases, consistent with the findings in UKBB. Shown are rg estimates and its 95% C.I. for diseases with a significant heritability for both traits.

To make a closer comparison between UKBB and FinnGen, we focused on the 26 mapped disease definitions. Most of these conditions had a higher in-sample prevalence in FinnGen compared to UKBB, except for high cholesterol (Table S19). Median age of first occurrence of FinnGen endpoints generally fell between UKBB-SR and the other two UKBB datasets. Locus-level comparison of age-of-first-occurrence GWAS revealed variable concordance between the two cohorts, with a few FinnGen traits showing a consistent direction of effects with UKBB with a sign-test p-value < 0.05 (e.g., hypertension, diabetes; Table S20; Figure S18). Heritability estimates showed that several of these diseases had a higher 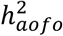 in FinnGen than in UKBB (e.g., hypothyroidism, UKBB <5.5%, FinnGen 8.0%; musculoskeletal disorders, UKBB ∼1.8%, FinnGen 4.7%; Figures 3B&S18; Table S19). Hypertension remained as the most significant trait with a non-zero 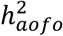, whereas angina and depression both showed little evidence of 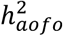 in either cohort. The significant 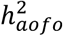 for asthma and high cholesterol in UKBB (SR and COMB) was not seen in FinnGen. These heterogeneous results for individual diseases might result from differences in sample ascertainment, criteria to define event occurrence age, as well as the sample size limitation of a case-only analysis.

Aside from these differences, genetic correlation analysis found that the majority of the heritable traits in FinnGen had a negative *r*_*g*_ between age of first occurrence and susceptibility (Figure 3C; Table S18), corroborating the UKBB finding of an inverse genomic relationship between event onset age and polygenic burden of the disease.

### MTAG of susceptibility and age of first occurrence improved polygenic risk prediction

Given the strong genetic overlap between susceptibility and age of first occurrence, we then performed Multi-trait Analysis of GWAS (MTAG)19 in UKBB to jointly analyze the two traits that shared a significant rg for each of the phenotypic datasets (Methods). For many of the tested disease endpoints, MTAG analysis identified additional significant loci not found in the original GWAS of susceptibility, showing an enhanced power for loci discovery equivalent to 0.5% to 30% increase in sample size. HIP endpoints had the least number of additional loci among the three datasets (Tables S21-23). The increase in association strength did not appear to occur uniformly across the genome, but rather, at loci associated with age of first occurrence or associated with specific life periods pertinent to a disease. For example, MTAG of asthma in the SR dataset re-captured some of the previously reported childhood-onset-specific loci not seen in GWAS, including a damaging missense variant on TESPA17 (12:55368291:C:T, MTAG P = 4.4×10-10; Figure 4A).

**Figure 4.**
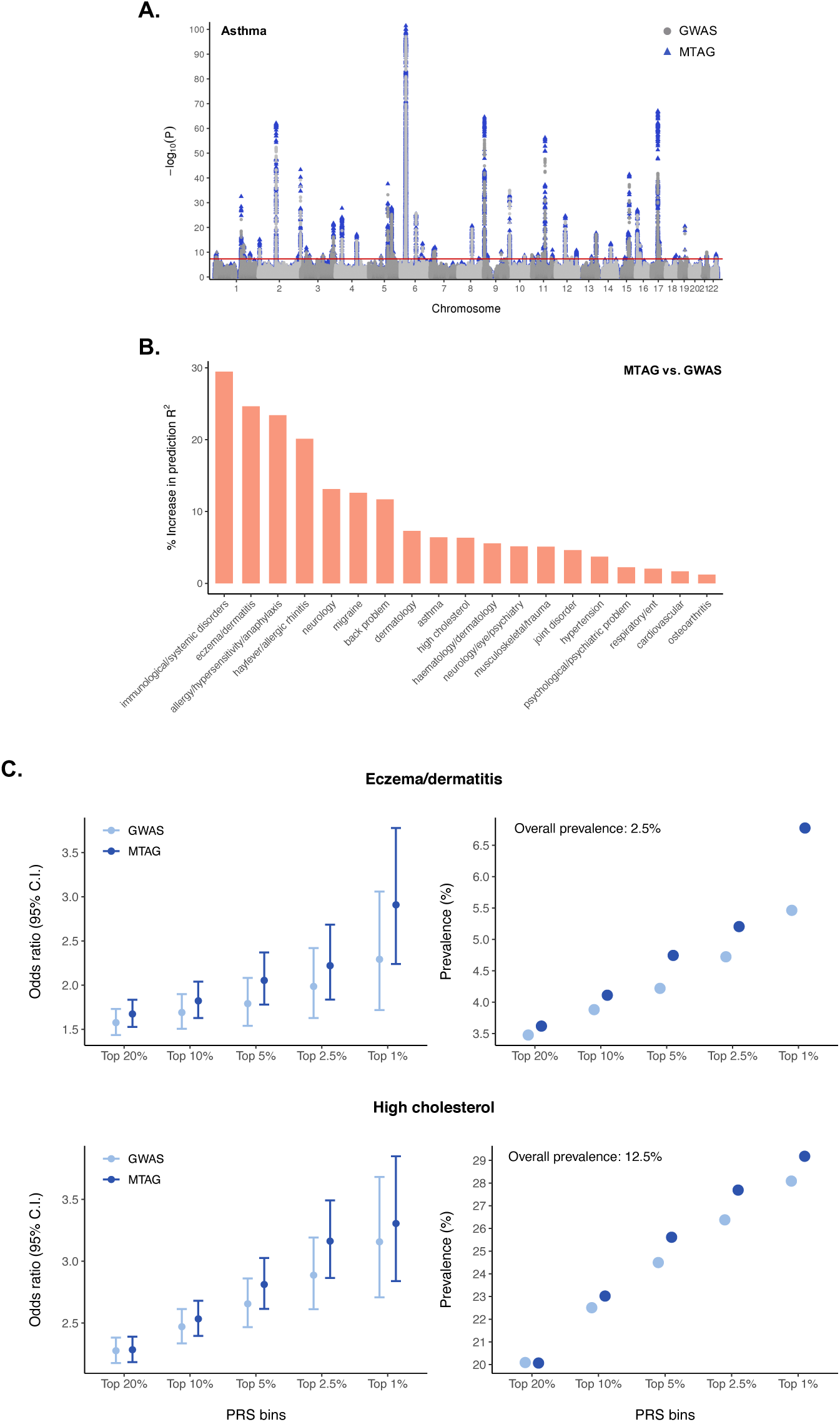
MTAG and PRS analysis in the UKBB SR dataset. **A**. A Manhattan plot of asthma MTAG that incorporates age of first occurrence information (blue triangle) compared to the original case-control GWAS (grey circle). The solid red line denotes P = 5×10-8. **B**. Improvement in disease risk prediction using MTAG-PRS versus GWAS-PRS. MTAG was performed for diseases with a significant *r*_*g*_ between age of first occurrence and susceptibility. PRS and the proportion increase in prediction R2 of MTAG relative to GWAS (y-axis) were computed in an independent sample of 91K EUR individuals. Results for the HIP and COMB datasets are shown in Figure S20. **C**. The application of MTAG-PRS and GWAS-PRS in risk stratification for two selected disease phenotypes. The left panel shows the adjusted odds ratio and its 95% CI (y-axis) comparing individuals in each of the top PRS percentiles (x-axis) to the rest of the population. Showing on the right is the corresponding disease prevalence in the top PRS percentiles computed using either GWAS or MTAG summary statistics. Full results for all three UKBB datasets can be found in Tables S24-26.

To explore how age of first occurrence could aid in risk prediction, we constructed polygenic risk score (PRS) using GWAS and MTAG summary statistics in the left-out set of 91,436 ancestry-matched individuals in UKBB. We calculated the relative change in prediction accuracy of MTAG versus GWAS on top of a covariate-only model (Methods). For diseases with an adequate sample size (>2000 cases in the target sample), the MTAG model that incorporated age of first occurrence outperformed the susceptibility-only GWAS in predicting disease risk for many traits. For SR conditions the increase in predictive accuracy can be as high as 30% and on average around 10%, while the improvement was overall less obvious for HIP endpoints. (Figures 4B&S20; Tables S24-26). Notably, diseases with a larger increase in prediction R^2^ seemed to involve several childhood-onset, allergy-related conditions (Figures 4B).

Finally, we evaluated the clinical utility of PRS in patient stratification using MTAG-PRS compared to GWAS-PRS (Methods). In both models, individuals in the top PRS percentiles (1%, 2.5%, 5%, 10%, and 20%) had a significantly elevated disease risk versus those who were not. Furthermore, for many traits, MTAG-PRS consistently identified individuals at a higher risk than predicted by GWAS, corresponding to a larger proportion of cases across top PRS percentiles (e.g., eczema, high cholesterol; Figures 4C&S21; Tables S24-S26). For instance, individuals in the top 1% GWAS-PRS were at a 2.2-fold increased risk for SR eczema compared to the average PRS group (P = 4.0×10-7) and a 2.3-fold risk than the rest of the sample (P = 1.7×10-8), while those defined by the top 1% MTAG-PRS had an even higher risk of OR = 2.5 (P = 1.9×10-11) and OR = 2.9 (P = 1.2×10-15), respectively (Figures 4C&S21). Overall, risk prediction and stratification incorporating age of first occurrence information showed more gains for SR phenotypes than defined in the other two datasets, particularly for conditions that tend to have a pediatric population.

## Discussion

Age of onset has long been an important phenotype of interest in epidemiological studies due to its rich information on timing of events relevant to disease etiology, but a systematic characterization of its genetic underpinnings has been lacking. Here, we provide a deep investigation into the genetic architecture of age of first occurrence of a wide range of complex disorders among White-British individuals in UKBB using three different phenotypic criteria (SR, HIP, COMB). We discovered that age of first occurrence has non-trivial genetic contributions across many diseases, some with unique genetic risk factors not associated with simply disease susceptibility. Although the extent of sharing varies by disease, genetic correlation analysis suggests that an earlier health-event occurrence indexes a heavier polygenic burden of the disease. Independent of UKBB, genetic investigation of the FinnGen cohort yields a similar pattern of heritability and genetic correlation estimates. Further, we demonstrate that incorporating onset age with susceptibility could improve risk loci discovery and patient stratification based on genetic risk prediction.

The scan of SNP-heritability of age of first occurrence 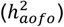 revealed a similar landscape of what has been observed with SNP-heritability of susceptibility to complex disorders 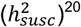. We note that 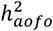 estimates the degree to which genetic variation explains age at first event *among* affected individuals whereas 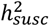 is defined at the population level and estimates the genetic contribution to a continuous disease liability. Notably, our use of different criteria to define age of onset of a disease and the inherent variability in the manifestation or diagnosis of a disease led to heterogeneous 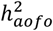 estimates between datasets, more so than variability among 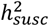 estimates (Figure S10). Aside from the sample size constraint, the extent to which 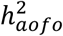 varied by source of definition is disease-dependent and may relate to health-reporting and health-seeking behaviors as well as the time span of each data type. The SR dataset serves as a good source for documenting the approximate age of onset for health events, including those starting early in life, but might suffer from recall bias, particularly for diseases with a mild manifestation.

On the other hand, for conditions that require in-patient stays, such as an acute onset (e.g., heart failure, diabetes complications), HIP phenotypes may be a more valid source. The downside of HIP-defined event occurrence is that it might not capture the exact age of onset for many diseases, including occurrences prior to when the ICD-10 system came into use and incorporated into the Health Episode Statistics database in the UK (1990s), and ICD codes can be given to patients for administrative purposes. The replication of our results in FinnGen, where registry data covers an older time period, partially addresses this limitation. Our analysis showed that SR and HIP endpoints often exhibit a differential pattern of prevalence and 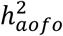 (Figure S11). The third approach of combining different sources to define the earliest event (e.g., UKBB-COMB and FinnGen) is most effective in increasing sample size, which is ideal for case-control GWAS but should be interpreted with caveats for age of onset GWAS. As different data sources can start at a different point in time and not all individuals are included in all datasets (e.g., Figure S16), this approach could result in a synthetic or multi-modal distribution of disease onset age not reflective of the actual timing of events. In short, there is unlikely to be one simple measure to define age of onset, and the heterogeneity in case ascertainment methods can affect the findings and interpretation of genetic analysis with respect to time.

Our findings suggest that the genetic basis of age of first occurrence and susceptibility— two seemingly orthogonal phenotypic components of a disease—have a correlated nature for many complex disorders. The shared and distinct loci between age of first occurrence and susceptibility indicate that some variants affect both disease risk and disease onset age while others are specifically associated with delaying or accelerating disease occurrence. The growth of biobank-scale datasets may reveal more genetic risk factors that can modify age of onset and may represent new therapeutic targets. On the genome-wide scale, our observation that earlier onset is associated with a greater polygenic loading for disease is consistent with what has long been hypothesized for polygenic disorders and has been increasingly observed in individual GWAS studies^13,14,21^. For Mendelian disorders, studies have also shown that its polygenic background can modify the age of disease occurrence among those with rare monogenic mutations^22,23^. Through genetic correlation analysis in the UKBB and FinnGen cohorts, we demonstrated that such an inverse genomic relationship between when and whether a disease occurs is in fact widespread among complex disorders. While the interpretation will vary by disease, a negative *r*_*g*_ might imply a heterogeneous genetic architecture at different ages of onset, suggesting age-related disease subtypes or a continuum whereby earlier onset of the disease is more genetically driven while later onset might reflect a greater contribution of non-genetic life events.

As an alternative to our case-only approach, Cox proportional hazard modeling, a common method to study time-to-event endpoints that models both disease status (whether) and onset time (when) accounting for censoring events, can be a powerful approach for detecting SNP-disease associations^24,25^. However, such models do not allow explicit assessment of genetic risk factors underlying age of first occurrence separately from disease susceptibility, and hence estimation of their genetic overlap. In addition, heritability from the Cox model is typically defined for cumulative hazards^25,26^ and does not have the same interpretation as 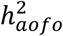. As more scalable survival models for GWAS are being developed^25,27^, a comparison of the two approaches will be informative for distinguishing their genetic findings and implications for the dissecting genetic architecture.

Through PRS analysis, we showed that genetic variation underlying age of first occurrence of health events can improve disease risk prediction and patient stratification, especially for diseases that tend to have an early onset. This demonstrates how axes of information correlated with case-control status can be useful in planning prevention and intervention strategies tailored toward individual genetic predisposition. The rich spectrum of phenotypic information in large biobank cohorts provides an unparalleled opportunity for epidemiological and genetic research to study clinical features beyond simply disease susceptibility through longitudinal healthcare records. Genome-wide analysis of such quantitative disease dimensions is critical and will continue to generate valuable insights into the genetic basis of disease development, severity, and progression.

## Online Methods

### UK Biobank, genotyping, and quality control

UK Biobank (UKBB) is a population-based cohort with extensive phenotype and genomic data on more than 500,000 individuals in the United Kingdom aged 40 to 69 years at recruitment^15,16^. The study started recruitment between 2006 and 2010 and has been followed up prospectively. The genetic data in the UK Biobank is available for a subset of 488,377 participants, with 49,950 individuals genotyped using the UK BiLEVE Axiom Array and the other 438,427 participants genotyped using the UK Biobank Axiom Array in GRCh37 coordinates. The Haplotype Reference Consortium (HRC)^28^ data and the merged UK10K and 1000 Genomes phase 3 data^29^ were used as reference panels for imputation.

We obtained the lists of post-QC samples and variants from the Neale Lab GWAS for analysis, which comprised 361,194 unrelated individuals of predominantly White-British descent and 13.7 million genetic markers. Quality control procedures of the genotype data were detailed in the Neale Lab blog posts and GitHub repository^20,30^. In brief, the initial UKBB cohort was filtered to those who were unrelated and did not have sex chromosome aneuploidy using the provided UKBB sample QC metrics. Among them, individuals who were self-reported “White-British”, “Irish”, or “White”, and fell within seven standard deviations of the first six principal components (PCs) were retained. Variants were filtered to those with an imputation INFO score > 0.8, a minor allele frequency (MAF) > 0.1% and a p-value for the Hardy-Weinberg equilibrium test > 1×10^−10^.

Here, we further removed participants who have since withdrawn consent and variants that were located on the sex chromosomes or had a MAF < 1%. The final dataset consisted of 361,140 individuals and 9.4 million autosomal, common variants. We then converted the data from .bgen to .pgen format using PLINK 2.0^31^, which preserved dosage information for association analysis.

### Age of first occurrence: phenotype definition and selection

Age of first occurrence of a disease or medical condition was estimated from three different phenotypic datasets in the UKBB for the 360K White-British subset, including *self-reported* medical conditions (SR), *hospital in-patient* records (HIP), and the *combined* first occurrence data-fields of diagnostic codes that mapped across different phenotypic datasets (COMB). Although the classification schemes are different, both SR and HIP datasets follow a tree-structure topology to define disease endpoints.

For the SR dataset, we aggregated all parent nodes and their children nodes from medical conditions ascertained through touchscreen questionnaires and verbal interviews (data-fields 20001 and 20002) into a total of 562 clinical terms across 11 non-cancer disease classes and 106 terms across 9 cancer categories. For each SR medical condition, participants were given the option to report either year or age when first diagnosed by a doctor in integers and the value was rounded to the nearest quarter age (data-fields 20007 and 20009). We identified affected individuals for each term and extracted their interpolated *age of diagnosis*. The earliest age of diagnosis among all children nodes was considered when the trait of interest was a parent node. Individuals who had a missing age of diagnosis—either uncertain/unknown or preferred not to answer—were excluded from the analysis.

The HIP dataset was based on the Health Episode Statistics database in the UK and contained hospitalization episodes for each participant in the form of International Classification of Diseases (ICD) classifications, predominantly in ICD-10. Each in-patient episode had a corresponding primary diagnosis and might be associated with one or more secondary diagnoses; dates of admission, episode-start, episode-end, and discharge were also recorded. The earliest dates in HIP can be traced back to 1990s, roughly when the ICD-10 system came into place (https://biobank.ctsu.ox.ac.uk/showcase/exinfo.cgi?src=Data_providers_and_dates). We mapped the available ICD-10 codes to PheWAS Codes (PheCodes)^32,33^ and defined all possible PheCode-based phenotypes based on its hierarchical structure, ranging from individual PheCodes, comorbid medical conditions with related PheCodes, to the entire PheCode category. Together, this resulted in 1,843 cleanly defined clinical terms across 18 PheCode categories. For each term, individuals who had at least one associated ICD-10 codes in the primary or secondary diagnoses were considered a case, and their *age of first hospitalization episode, or in-patient diagnosis*, was calculated as the interval between the earliest episode-start date and the mid-month of their birth year. Where episode-start date was missing, admission date was used instead.

The COMB dataset is a special data type that contains the first-reported dates of a diagnostic code for a range of different non-cancer health outcomes, generated by mapping across SR, HIP, primary care, and death records. Note that the current release of primary care data includes only 45% of the UKBB cohort. For data types not based on the ICD-10 classification system, health outcomes were first mapped to a related 3-character ICD-10 code where appropriate by the UKBB team, and its date of first occurrence was recorded as the earliest among the four different datasets. The mapping mechanism however has not been externally validated and many of the SR endpoints did not have a corresponding ICD-10 code. We extracted all the available first occurrence fields, totaling 1,165 terms across 16 disease categories (data-fields 2401-2417), set to missing any improbable dates for each term (e.g., an event date before birth, at birth, or in the future), and estimated the number of affected individuals as those with a non-missing first occurrence date. Due to the composite nature of COMB endpoints, we did not map these 3-character ICD-10 codes to PheCodes. Age when a given condition was first reported was estimated as described for the HIP dataset.

We noted that age first occurrence estimated in the SR and the COMB datasets could be as early as <1 year of age, and the small values seemed unlikely for some diseases. However, such instances only accounted for a tiny fraction of the total affected individuals, particularly among adult-onset conditions (Tables S4&S6). Imposing any threshold to truncate the distribution would seem arbitrary given that the biologically plausible age of onset differs widely across diseases. We instead acknowledged that measurement error or misclassification may exist for these small values but their impact on analysis should be limited as a whole.

To compare the distribution of age of first occurrence across data types, we selected 26 phenotypes whose definitions could be well mapped between SR and HIP, 15 of which were comparable across all three datasets (hypertension, angina, myocardial infarction, heart arrhythmia, hypothyroidism, high cholesterol, diabetes, asthma, depression, migraine, osteoarthritis, disc problem, gastroesophageal reflux disease/GERD, hiatus hernia, cholelithiasis/gallstones, breast cancer, skin cancer, endocrine/metabolic disorders, psychiatric disorders, neurological diseases, cardiovascular diseases, respiratory diseases, gastrointestinal/digestive diseases, dermatologic diseases, musculoskeletal disorders, and neoplasms) (Table S7).

### Association analysis

We performed a GWAS of age of first occurrence for diseases with at least 5000 affected individuals, a cutoff we considered sufficiently powered for genetic analysis. This led to a total of 70 SR, 224 HIP, and 164 COMB medical conditions (Tables S1-3). Age of first occurrence was analyzed as a quantitative outcome in a linear regression to estimate its association with imputed SNP dosages in PLINK 2.0 (PLINK v2.00a2LM), adjusting for sex, genotyping array, and the first 20 PCs. For comparison, a GWAS of susceptibility to the same disease definition was also performed, which treated “ever affected with the condition” as the endpoint and considered all non-case individuals as controls; the same sets of covariates were included in the regression model. We deliberately did not adjust for current age in the model to avoid double counting the age information. Nonetheless, when current age was included as an additional covariate, we observed comparable GWAS results and heritability estimates.

To obtain independently associated loci, we performed linkage disequilibrium (LD) clumping on GWAS summary statistics. The clumping procedure started by identifying genome-wide significant SNPs (p-value < 5×10^−8^) and then selecting any other SNPs that had a r^2^ > 0.1 with the index SNP within a 500kb window to form a clump (--clump-p1 5e-08 --clump-p2 0.05 -- clump-r2 0.1 --clump-kb 500). The procedure stopped when all genome-wide significant SNPs were assigned to a locus. Any overlapping associated loci were merged using BEDTools/bedtools^34^ (v2.27.1). A randomly selected sample of 10,000 White British individuals in UKBB was used as the reference panel to compute LD. Due to its strong and extensive LD structure, the major histocompatibility complex (MHC) region (chr6: 25Mb-35Mb) was treated as one genomic locus. Finally, associated genomic loci of age of first occurrence and susceptibility from their respective GWAS were compared using “bedtools intersect” to identify shared or distinct loci.

### Inflation evaluation, SNP-heritability, and genetic correlation

We performed LD Score regression (LDSR)^35^ analysis on GWAS summary statistics to assess the extent of residual confounding and to estimate SNP-heritability. LDSR can distinguish inflation in GWAS association ͼ^2^ statistics due to confounding such as population stratification from true polygenicity. An LDSR intercept close to one, or a ratio of (intercept-1)/(mean ͼ^2^−1) close to zero, would indicate the contribution of confounding biases is well-controlled. The analysis was done using the pre-computed LD scores of 1.2 million high-quality HapMap3 SNPs (excluding the MHC region) from the European samples in the 1000 Genomes Project^36^ and GWAS summary-level results from the tested UKBB disease phenotypes. SNP-heritability for age of first occurrence was estimated based on the slope of LDSR to measure the degree to which the phenotypic variation is explained by common genetic variation. SNP-heritability for susceptibility was estimated on the liability-scale, assuming that population prevalence equals sample prevalence in the dataset. For conditions with a significant heritability of its age of first occurrence and susceptibility endpoints, we calculated a genetic correlation (*r*_*g*_) between the two traits using bivariate LDSR^37^. The genetic covariance is estimated using the slope from the regression of the product of z-scores from two GWAS studies against the LD score. The estimate obtained from this method represents the genetic correlation between the two traits attributable to all polygenic effects captured by common SNPs. In addition, we estimated pairwise *r*_*g*_’s separately for age-of-first-occurrence and susceptibility phenotypes to examine the pattern of genetic sharing across diseases.

### Replication analysis in FinnGen

To replicate the results of the heritability pattern of age of first occurrence and its genetic correlation with susceptibility, we analyzed the FinnGen cohort^17^ of 130,423 unrelated individuals. Age at recruitment of FinnGen participants ranged from 0.08 to 98.98 (median: 54.36, IQR: 25.45). FinnGen is a public-private partnership project combining genotype data from Finnish biobanks and digital health record data from Finnish health registries (https://www.finngen.fi/en). Six regional and three country-wide Finnish biobanks participate in FinnGen, which also includes data from previously established populations and disease-based cohorts.

FinnGen disease endpoints are defined using nationwide registries. Data are harmonized over the ICD revisions 8, 9 and 10, cancer-specific ICD-O-3, (NOMESCO) procedure codes, Finnish-specific Social Insurance Institute (KELA) drug reimbursement codes, and ATC-codes for medications. These registries span decades (Figure S16) and are electronically linked to the cohort baseline data using the unique national personal identification numbers assigned to all Finnish citizens and residents. We used genotype and phenotype data from FinnGen release v4 of 130,423 unrelated Finnish participants, excluding population outliers via PCA and related individuals (<3rd degree) using the KING software^38^ (Supplementary Notes).

We analyzed diseases with at least 5000 cases and a few additional ones with slightly fewer than 5000 cases that we considered relevant for comparison with UKBB, leading to a total of 280 medical conditions. For each condition, age of first occurrence was defined as the earliest age of an event in the registries. Age of first occurrence across the analyzed diseases ranged from 0 to 105.65 (median: 52.04, IQR: 28.22). fastGWA^39^ linear regression model was used for GWAS analysis. Sex, 10 PCs, and genotyping batch were used as covariates. Low quality variants with missingness rate > 0.1 and variants with MAF < 0.0001 were excluded from the analysis. Following the same procedures in the UKBB analysis, we identified loci uniquely associated with age of first occurrence, estimated the SNP-heritability of age of first occurrence and susceptibility using LDSR, and computed a genetic correlation between them for each analyzed disease definition. Standard LD scores were used based on the 1000 genomes reference set, restricting to European populations.

In addition to the phenome-wide analysis, we focused on the 26 mapped phenotypes to compare the results in UKBB and FinnGen (Table S19). To evaluate how the estimated effect sizes of age of first occurrence in UKBB replicate in FinnGen, we first identified the SNPs present both in UKBB and FinnGen. For the common SNPs, we then obtained the independently associated loci in the UKBB performing LD clumping with a p-value threshold of 0.0001. We finally looked at the correlation of the effect sizes in UKBB and FinnGen for the index SNPs of each clump and evaluated the concordance of the direction of the effects with a one-sample binomial test.

### MTAG analysis of susceptibility and age of first occurrence

For UKBB diseases where age of first occurrence and susceptibility shared a significant genetic correlation, we meta-analyzed the two traits using Multi-Trait Analysis of GWAS (MTAG)^19^. Built upon the LDSR framework, MTAG boosts power for loci discovery for a trait by factoring in its shared genetic architecture with other traits while accounting for sample overlap. MTAG by default imposes additional SNP filters, which resulted in ∼7.8 million SNPs, fewer than included in the GWAS. All the comparisons of MTAG and GWAS were based on this subset of SNPs. The number of significant loci in MTAG of *susceptibility*—now with the information on age of first occurrence incorporated—was calculated based on the same clumping and merging procedure described previously and were compared against the original susceptibility GWAS to identify any additional loci.

### Polygenic risk prediction of disease susceptibility

To evaluate if genetic sharing with age of first occurrence can improve risk prediction of disease susceptibility, we constructed polygenic risk scores (PRS) from MTAG association statistics and compared its performance to that based on the susceptibility GWAS. We used PRS-CS^40^, a Bayesian polygenic prediction method that imposes a continuous shrinkage prior on SNP effect sizes and is robust to diverse genetic architectures, to obtain the posterior effect size of each SNP, using 1000 Genomes European samples as the LD reference panel. PRS was calculated as the sum of allele dosages weighted by the posterior effect sizes of each SNP in an independent, ancestry-matched sample of 91,436 UKBB Individuals using PLINK 2.0 (--score). The target sample was selected as follows: starting with 1000 Genomes variants that overlapped with UKBB genotyped variants, we filtered to high-quality autosomal SNPs (no strand-ambiguous alleles, not in long-range LD regions, with a call rate > 0.98 and a MAF > 0.05), and pruned for LD (r^2^ < 0.2) down to 149,501 nearly independent SNPs. Using the pruned SNPs, we performed PCA on the 2,504 individuals in 1000 Genomes data and then projected the 488,377 UKBB individuals onto the computed PC space. With the first 6 PCs in 1000 Genomes as the training data, we used the Random Forest classifier to assign a “super population” label with a prediction probability ≥ 0.9 for each UKBB participant (AFR, AMR, EAS, EUR, or SAS). This resulted in 91,436 individuals who were classified as EUR and were not included in the discovery GWAS. We focused on diseases with >2,000 cases in the target sample for PRS analysis.

To estimate the predictive power of PRS, we calculated an incremental McFadden’s pseudo-R^2^ (*pR*^2^), comparing a logistic regression model that included PRS and a set of covariates (age, sex, genotyping array, and the top 10 PCs) to a covariate-only model. The percentage improvement in the predictive power of MTAG-PRS over GWAS-PRS was computed as 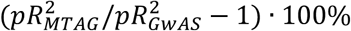· 100%. Next, we evaluated the performance of risk stratification of MTAG-PRS and GWAS-PRS; both PRS were standardized to have a mean of 0 and a standard deviation of 1. We dichotomized individuals into those who belonged to the top PRS percentile (1%, 2.5%, 5%, 10%, and 20%) versus those who did not, or those among the average percentiles (20-80%). We then modeled disease risk as a function of the binary PRS indicator as well as age, sex, genotyping array, and the top 10 PCs as covariates. The proportion of cases (prevalence) was also identified in each top PRS percentile.

## Data Availability

The UK Biobank data may be obtained through online registration and application (https://www.ukbiobank.ac.uk/). Study details of the FinnGen study cohort may be accessed through Finnish Biobanks' FinnBB portal (www.finbb.fi). GWAS summary statistics of age of first occurrence generated in the study will be made available at a public data repository prior to publication.

## Acknowledgements

We thank Chia-Yen Chen and Raymond Walters for their insightful suggestions and comments. We would like to acknowledge the participants and investigators of the UK Biobank and the FinnGen study for their gracious contribution. This research has been conducted using the UK Biobank Resource under Application Numbers 31063 and 32568. T.G. is supported by the National Institutes of Health (NIH) grant K99/R00 AG054573. A special thanks to the Neale Lab UKBB team (Liam Abbott, Sam Bryant, Claire Churchhouse, Andrea Ganna, Daniel Howrigan, Duncan Palmer, Ben Neale, Raymond Walters, Caitlin Carey, Cotton Seed, Jonathan Bloom, Tim Poterba, Dan King, Jackie Goldstein, Arcturus Wang, Patrick Schultz, John Compitello, and Jack Goldsmith) who together contributed to data management, quality control, and analysis of the UKBB genotype data and made QC metrics and results publicly available to the scientific community.

## Author contribution

Y.A.F. conceived the idea of the project through discussions and guidance from T.G., J.W.S. and B.M.N. Y.A.F. conducted the primary analysis and drafted the manuscript with feedback from all co-authors. T.G. provided logistic and methodological support for the UK Biobank analysis. M.C. contributed to FinnGen analysis and writing methods, with A.G. overseeing the analysis. J.W.S. and B.M.N. co-supervised the work and provided critical interpretation of the results.

## References

1. Claussnitzer, M. et al. A brief history of human disease genetics. Nature 577, 179–189 (2020).

2. Visscher, P.M. et al. 10 Years of GWAS Discovery: Biology, Function, and Translation. Am J Hum Genet 101, 5–22 (2017).

3. Kamboh, M.I. et al. Genome-wide association analysis of age-at-onset in Alzheimer’s disease. Mol Psychiatry 17, 1340–6 (2012).

4. Woolston, A.L. et al. Genetic loci associated with an earlier age at onset in multiplex schizophrenia. Scientific Reports 7, 6486 (2017).

5. Blauwendraat, C. et al. Parkinson’s disease age at onset genome-wide association study: Defining heritability, genetic loci, and α-synuclein mechanisms. Mov Disord 34, 866–875 (2019).

6. Ferreira, M.A.R. et al. Genetic Architectures of Childhood- and Adult-Onset Asthma Are Partly Distinct. Am J Hum Genet 104, 665–684 (2019).

7. Pividori, M., Schoettler, N., Nicolae, D.L., Ober, C. & Im, H.K. Shared and distinct genetic risk factors for childhood-onset and adult-onset asthma: genome-wide and transcriptome-wide studies. Lancet Respir Med 7, 509–522 (2019).

8. Power, R.A. et al. Genome-wide Association for Major Depression Through Age at Onset Stratification: Major Depressive Disorder Working Group of the Psychiatric Genomics Consortium. Biol Psychiatry 81, 325–335 (2017).

9. Ge, T., Chen, C.Y., Neale, B.M., Sabuncu, M.R. & Smoller, J.W. Phenome-wide heritability analysis of the UK Biobank. PLoS Genet 13, e1006711 (2017).

10. Davis, O.S.P., Haworth, C.M.A. & Plomin, R. Dramatic Increase in Heritability of Cognitive Development from Early to Middle Childhood: An 8-Year Longitudinal Study of 8,700 Pairs of Twins. Psychological Science 20, 1301–1308 (2009).

11. Nivard, M.G. et al. Stability in symptoms of anxiety and depression as a function of genotype and environment: a longitudinal twin study from ages 3 to 63 years. Psychological Medicine 45, 1039–1049 (2015).

12. Gottesman, I. & Shields, J. A polygenic theory of schizophrenia. Proc Natl Acad Sci U S A 58, 199–205 (1967).

13. Musliner, K.L. et al. Association of Polygenic Liabilities for Major Depression, Bipolar Disorder, and Schizophrenia With Risk for Depression in the Danish Population. JAMA Psychiatry 76, 516–525 (2019).

14. Mars, N. et al. Polygenic and clinical risk scores and their impact on age at onset and prediction of cardiometabolic diseases and common cancers. Nat Med 26, 549–557 (2020).

15. Bycroft, C. et al. The UK Biobank resource with deep phenotyping and genomic data. Nature 562, 203–209 (2018).

16. Sudlow, C. et al. UK biobank: an open access resource for identifying the causes of a wide range of complex diseases of middle and old age. PLoS Med 12, e1001779 (2015).

17. FinnGen. FinnGen Documentation of R3 release (https://finngen.gitbook.io/documentation/). (2020).

18. Borodulin, K. et al. Cohort Profile: The National FINRISK Study. Int J Epidemiol 47, 696–696i (2018).

19. Turley, P. et al. Multi-trait analysis of genome-wide association summary statistics using MTAG. Nat Genet 50, 229–237 (2018).

20. http://www.nealelab.is/uk-biobank/.

21. Ferreira, M.A.R. et al. Age-of-onset information helps identify 76 genetic variants associated with allergic disease. PLoS Genet 16, e1008725 (2020).

22. Genetic Modifiers of Huntington’s Disease (GeM-HD) Consortium. Identification of Genetic Factors that Modify Clinical Onset of Huntington’s Disease. Cell 162, 516–26 (2015).

23. Fahed, A.C. et al. Polygenic background modifies penetrance of monogenic variants for tier 1 genomic conditions. Nature Communications 11, 3635 (2020).

24. Staley, J.R. et al. A comparison of Cox and logistic regression for use in genome-wide association studies of cohort and case-cohort design. Eur J Hum Genet 25, 854–862 (2017).

25. He, L. & Kulminski, A.M. Fast Algorithms for Conducting Large-Scale GWAS of Age-at-Onset Traits Using Cox Mixed-Effects Models. Genetics 215, 41–58 (2020).

26. Yazdi, M.H., Visscher, P.M., Ducrocq, V. & Thompson, R. Heritability, reliability of genetic evaluations and response to selection in proportional hazard models. J Dairy Sci 85, 1563–77 (2002).

27. Dey, R. et al. An efficient and accurate frailty model approach for genome-wide survival association analysis controlling for population structure and relatedness in large-scale biobanks. bioRxiv (2020).

28. McCarthy, S. et al. A reference panel of 64,976 haplotypes for genotype imputation. Nat Genet 48, 1279–83 (2016).

29. Huang, J. et al. Improved imputation of low-frequency and rare variants using the UK10K haplotype reference panel. Nat Commun 6, 8111 (2015).

30. https://github.com/Nealelab/UK_Biobank_GWAS.

31. Chang, C.C. et al. Second-generation PLINK: rising to the challenge of larger and richer datasets. Gigascience 4, 7 (2015).

32. Denny, J.C. et al. Systematic comparison of phenome-wide association study of electronic medical record data and genome-wide association study data. Nat Biotechnol 31, 1102–10 (2013).

33. Wu, P. et al. Mapping ICD-10 and ICD-10-CM Codes to Phecodes: Workflow Development and Initial Evaluation. JMIR Med Inform 7, e14325 (2019).

34. Quinlan, A.R. & Hall, I.M. BEDTools: a flexible suite of utilities for comparing genomic features. Bioinformatics 26, 841–2 (2010).

35. Bulik-Sullivan, B.K. et al. LD Score regression distinguishes confounding from polygenicity in genome-wide association studies. Nat Genet 47, 291–5 (2015).

36. 1000 Genomes Project Consortium et al. A global reference for human genetic variation. Nature 526, 68–74 (2015).

37. Bulik-Sullivan, B. et al. An atlas of genetic correlations across human diseases and traits. Nat Genet 47, 1236–41 (2015).

38. Manichaikul, A. et al. Robust relationship inference in genome-wide association studies. Bioinformatics 26, 2867–73 (2010).

39. Jiang, L. et al. A resource-efficient tool for mixed model association analysis of large-scale data. Nat Genet 51, 1749–1755 (2019).

40. Ge, T., Chen, C.Y., Ni, Y., Feng, Y.A. & Smoller, J.W. Polygenic prediction via Bayesian regression and continuous shrinkage priors. Nat Commun 10, 1776 (2019).

