## Supplementary Figures for "Findings and insights from the genetic investigation of age of first reported occurrence for complex disorders in the UK Biobank and FinnGen"

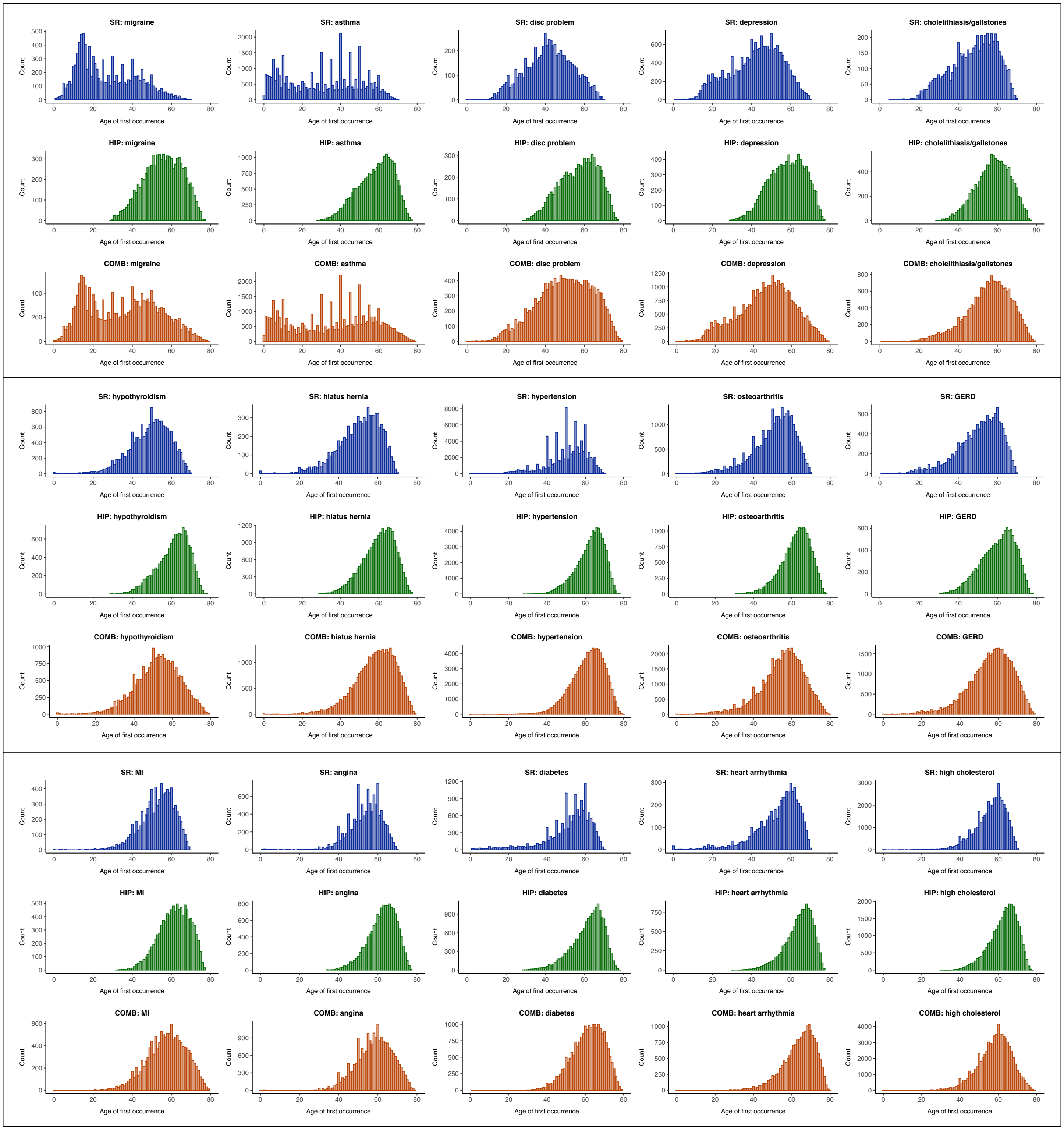


**Figure S1.** Age of first occurrence distribution for 15 disease phenotypes with comparable definitions across self-reported (SR; blue), hospital in-patient (HIP; green), and the combined first-occurrence (COMB; orange) datasets.

Distribution of age of first occurrence differs by trait and data source. Age of first occurrence typically ranges from 0-70 in the SR dataset, 30-80 in the HIP dataset, and 0-80 years of age in the COMB dataset. Comparison of an additional 11 phenotypes mapped only between SR and HIP is available in Table S7.

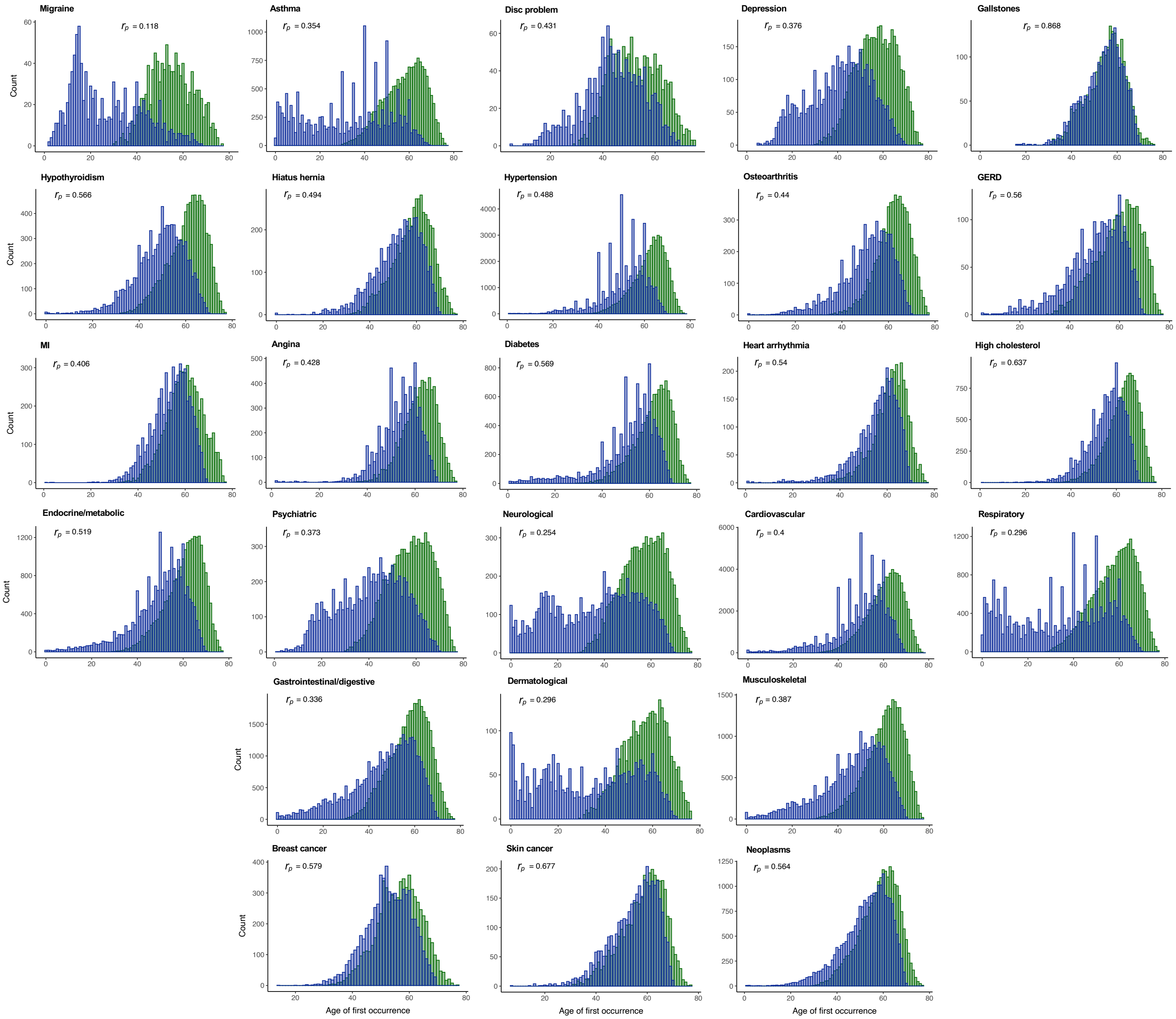


**Figure S2.** Age of first occurrence distribution of affected individuals recorded in both SR (blue) and HIP (green) datasets for the 26 mapped disease definitions.

A Pearson's correlation coefficient ($r_p$) was estimated to quantify the phenotypic similarity in age of first occurrence between the SR and HIP for each disease. The overlap between the two data sources varies from little to moderate, with an average $r_p$ of 0.46 across all 26 diseases.

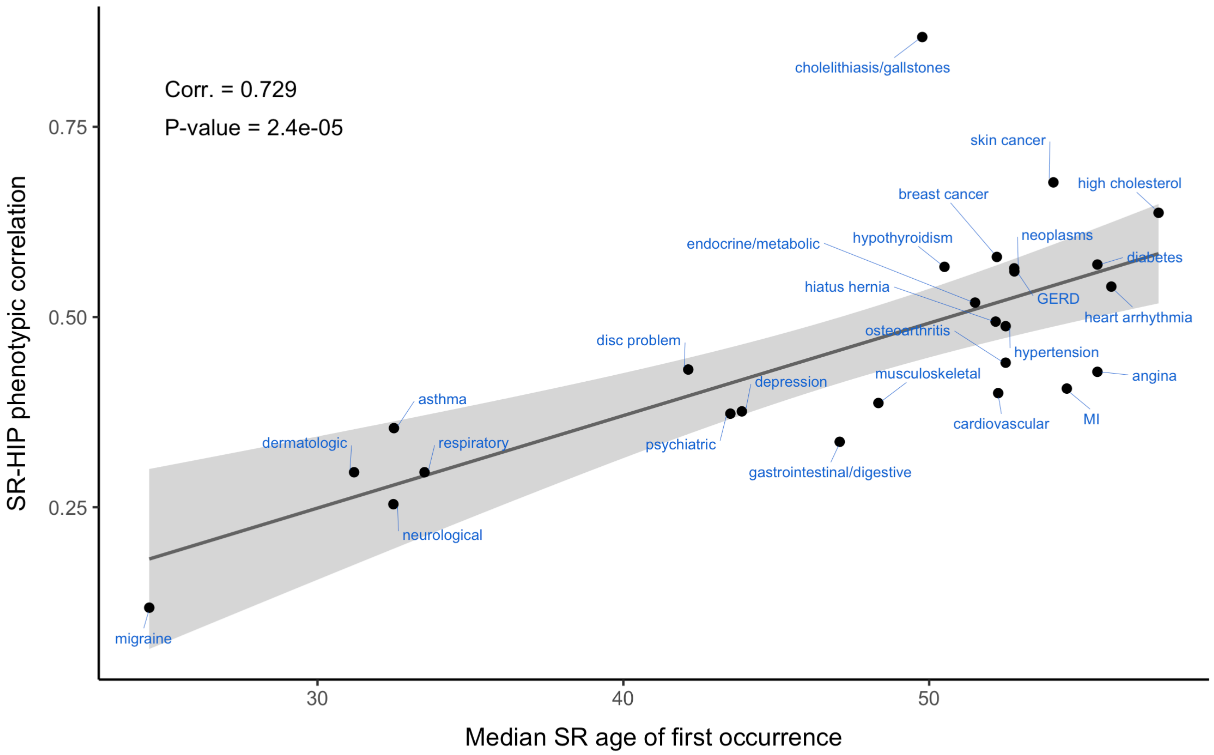


**Figure S3.** Phenotypic correlation ($r_p$) of age of first occurrence between SR and HIP endpoints increases with median age of first occurrence across 26 mapped diseases

Median age of first occurrence in the SR dataset is plotted against the cross-dataset Pearson's correlation coefficient between SR and HIP age of first occurrence ($r_p$) for these 26 mapped diseases. The solid grey line indicates the linear regression line estimated from the data, along with a 95% confidence band shown in the shaded gray area.

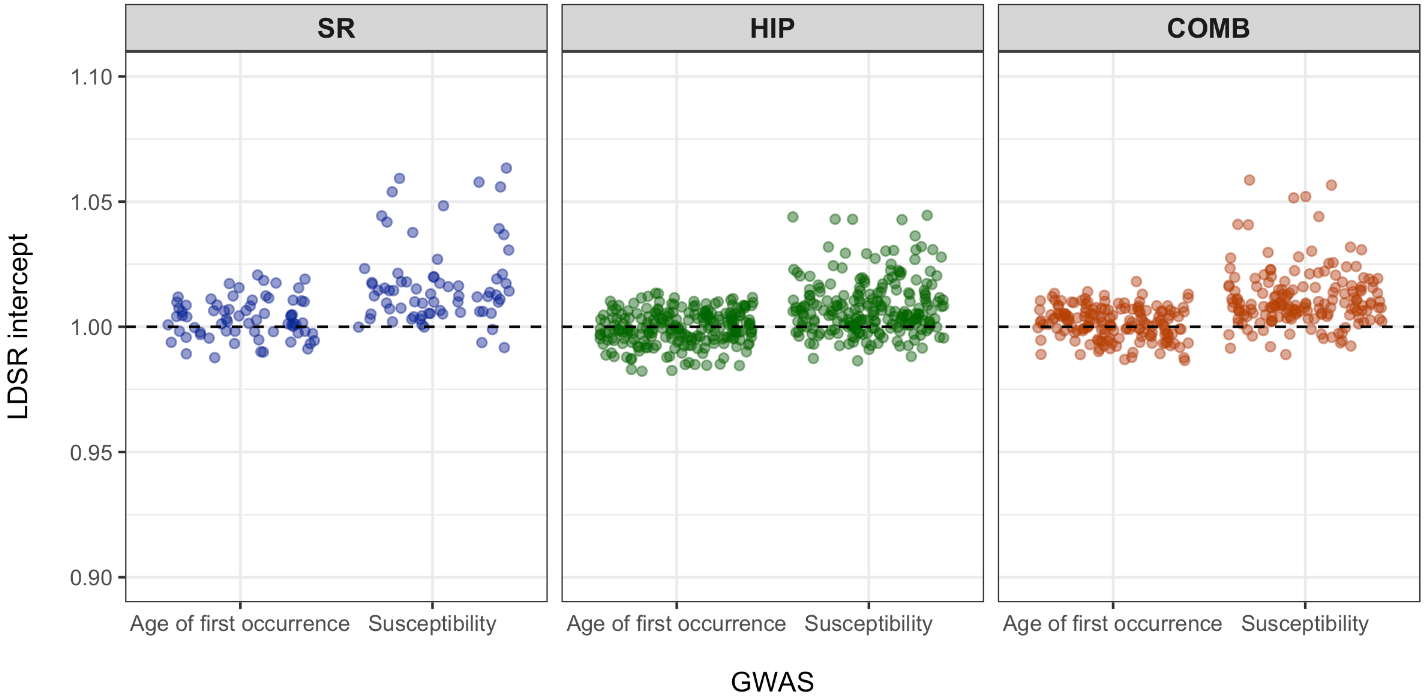


**Figure S4.** Univariate LDSR intercepts across all performed GWASes

A LD Score Regression (LDSR) intercept was estimated for the GWAS of 70 SR, 224 HIP, and 164 COMB disease definitions with >5000 affected individuals.

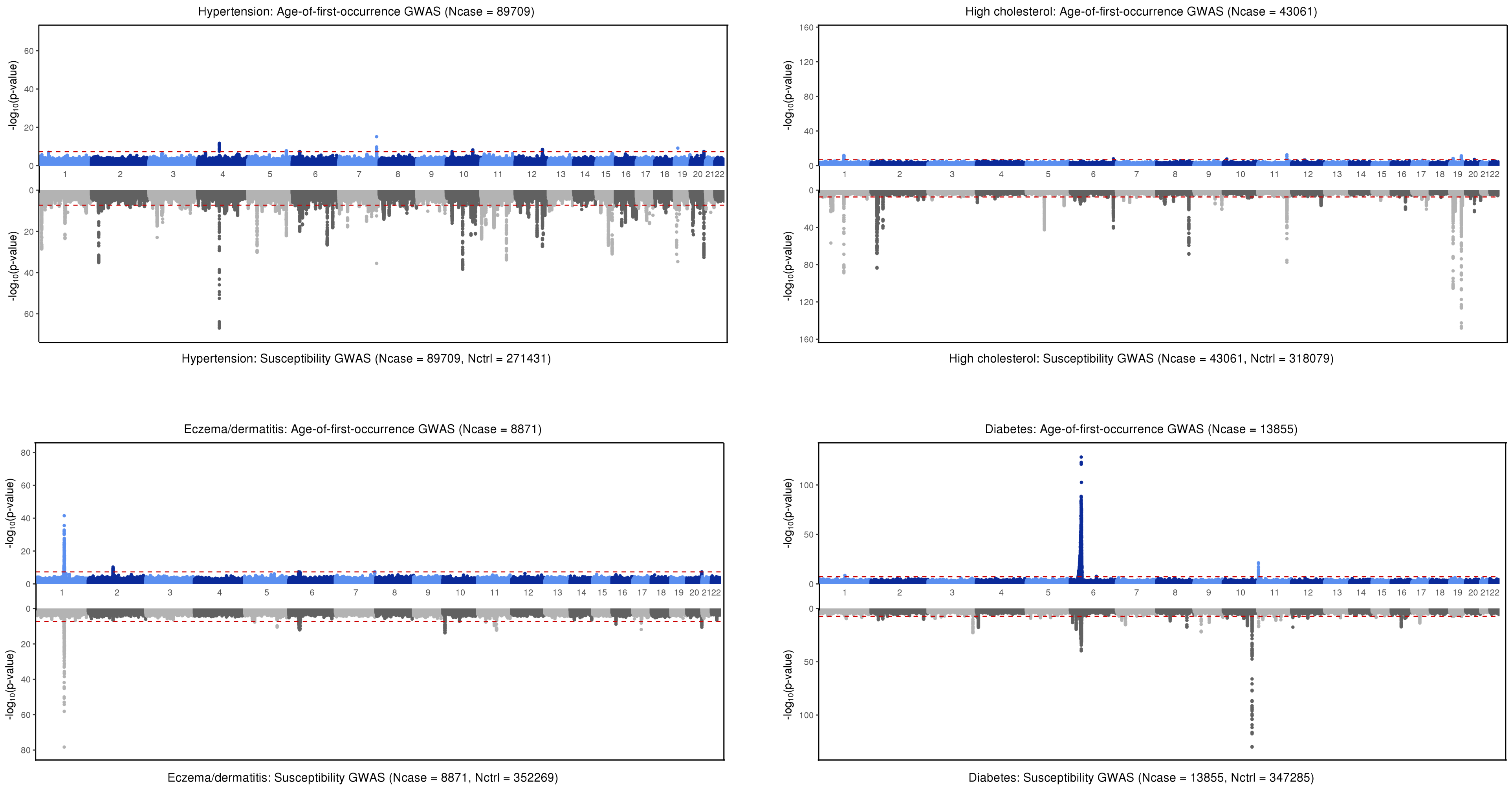


**Figure S5. Miami plots of GWAS results for selected disease endpoints in the SR dataset**

Each figure consists of a GWAS of age-of-first-occurrence (upper panel) and a GWAS of disease susceptibility (lower panel) for a given disease phenotype. P-values are shown on the $-\log_{10}$ scale on the y-axis, plotted against chromosome positions on the x-axis. The red dotted lines denote the genome-wide significance threshold at $P = 5{\times}10^{-8}$.

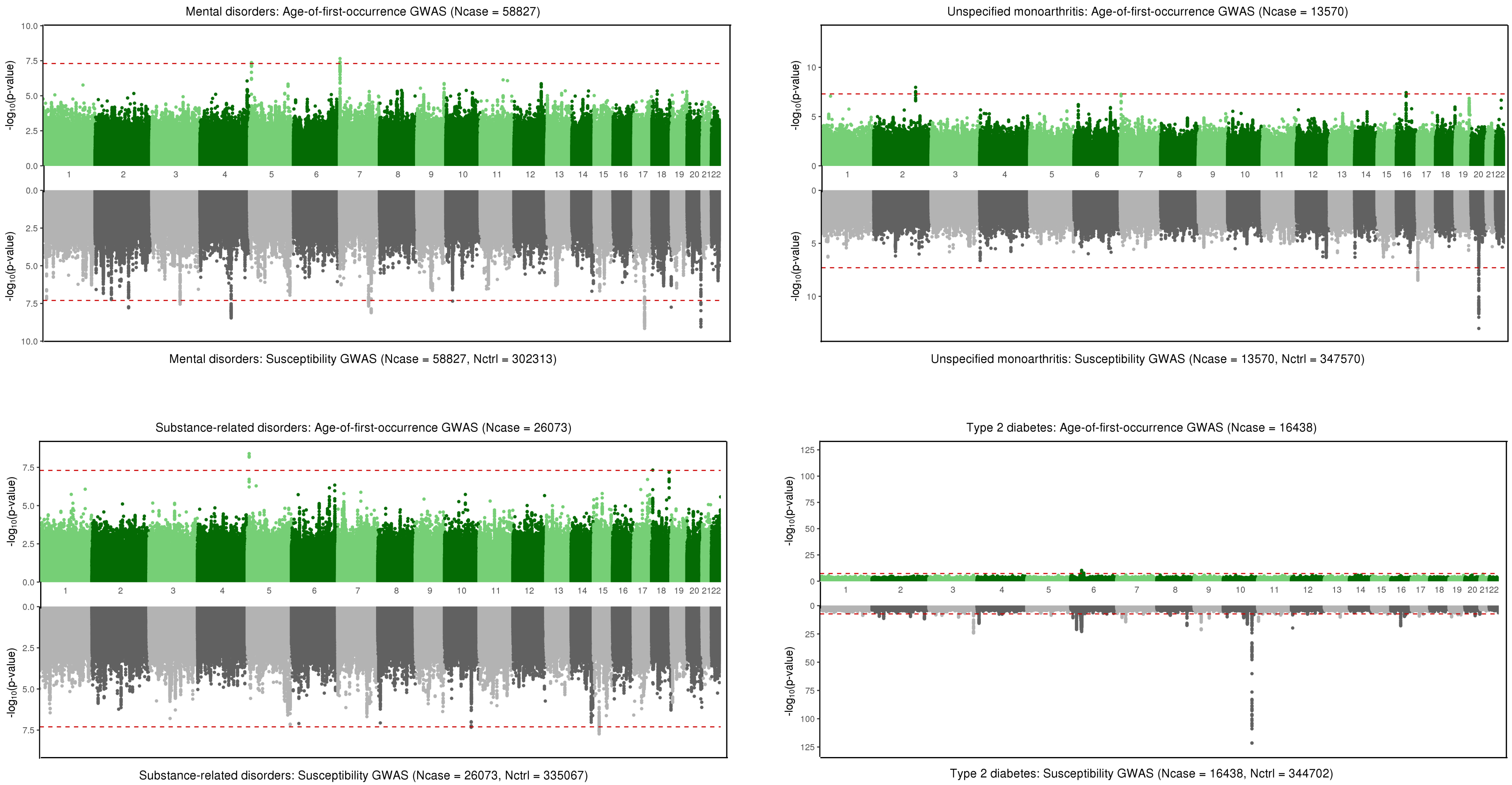


**Figure S6. Miami plots of GWAS results for selected disease endpoints in the HIP dataset**

Each figure consists of a GWAS of age-of-first-occurrence (upper panel) and a GWAS of disease susceptibility (lower panel) for a given disease phenotype. P-values are shown on the $-\log_{10}$ scale on the y-axis, plotted against chromosome positions on the x-axis. The red dotted lines denote the genome-wide significance threshold at $P = 5\times10^{-8}$.

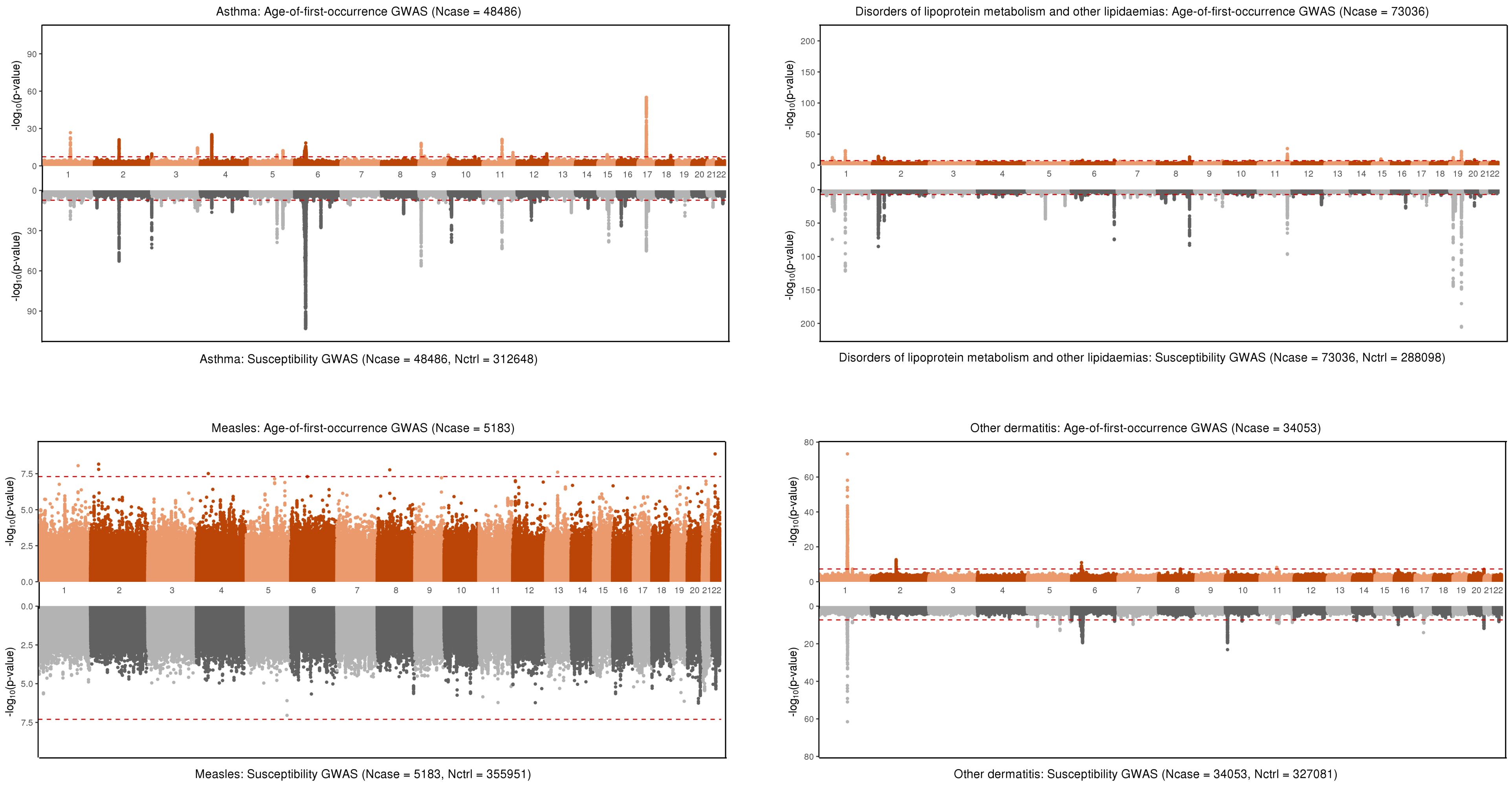


**Figure S7. Miami plots of GWAS results for selected disease endpoints in the COMB dataset**

Each figure consists of a GWAS of age-of-first-occurrence (upper panel) and a GWAS of disease susceptibility (lower panel) for a given disease phenotype. P-values are shown on the $-\log_{10}$ scale on the y-axis, plotted against chromosome positions on the x-axis. The red dotted lines denote the genome-wide significance threshold at $P = 5\times10^{-8}$.

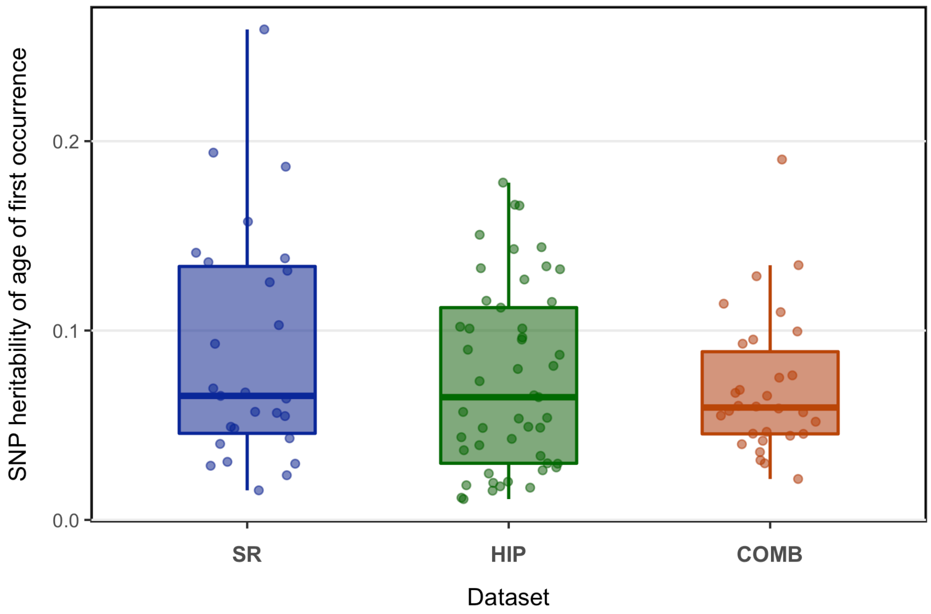


**Figure S8.** SNP-heritability of age-of-first-occurrence traits ($h^2_{aofo}$) with a nominal p-value < 0.05

$h^2_{aofo}$ was estimated using univariate LDSR for disease definitions with >5000 affected individuals. Shown here includes 27 SR endpoints, 49 HIP endpoints, and 30 COMB endpoints that have a significantly non-zero $h^2_{aofo}$. Each dot represents an individual disease.

**A.**

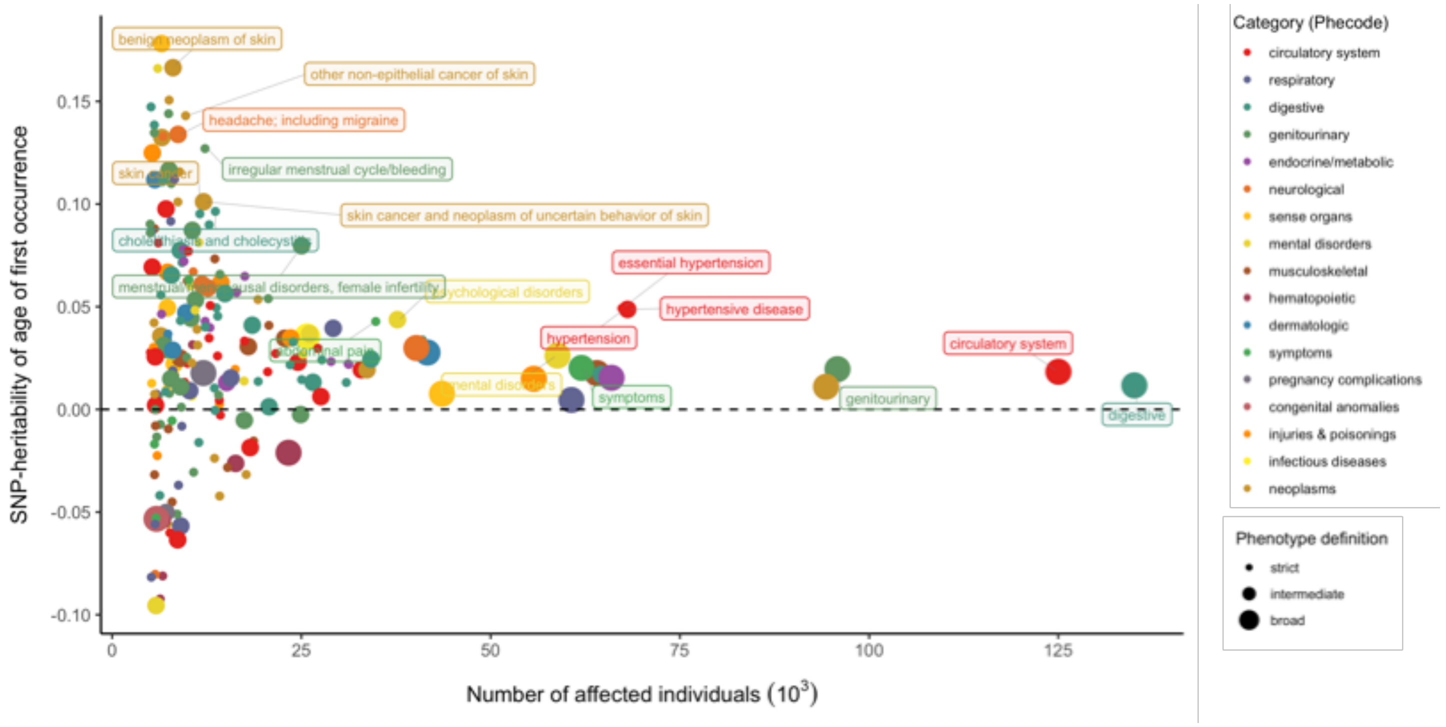


**B.**

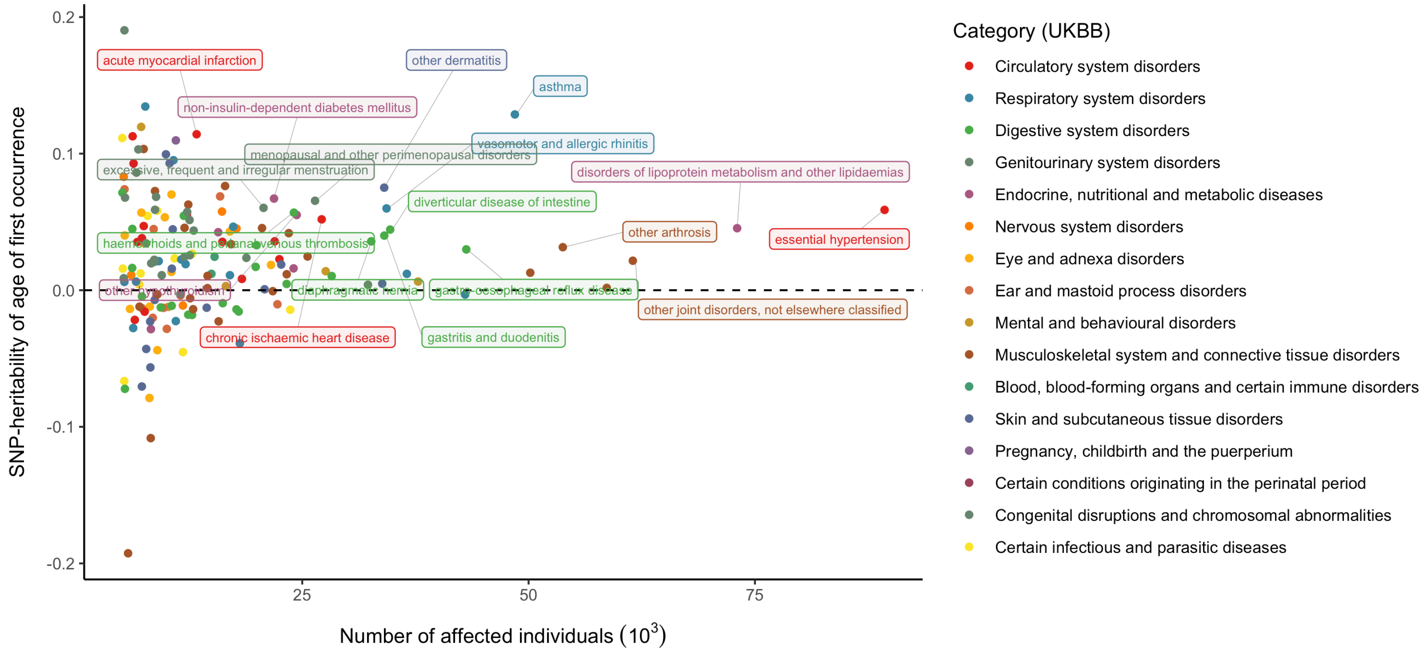


**Figure S9.** SNP-heritability estimates of age-of-first-occurrence endpoints ($h^2_{aofo}$) in the HIP dataset (**A**) and in the COMB dataset (**B**)

$h^2_{aofo}$ was estimated from univariate LDSR across 224 HIP and 164 COMB disease definitions, respectively. Each dot represents an individual disease, colored by disease categories used in each dataset; a larger dot corresponds to a broader disease definition. Labeled are conditions with a significant $h^2_{aofo}$ at FDR < 0.05. Heritability analysis of SR diseases reveal a similar pattern in Figure 3B.

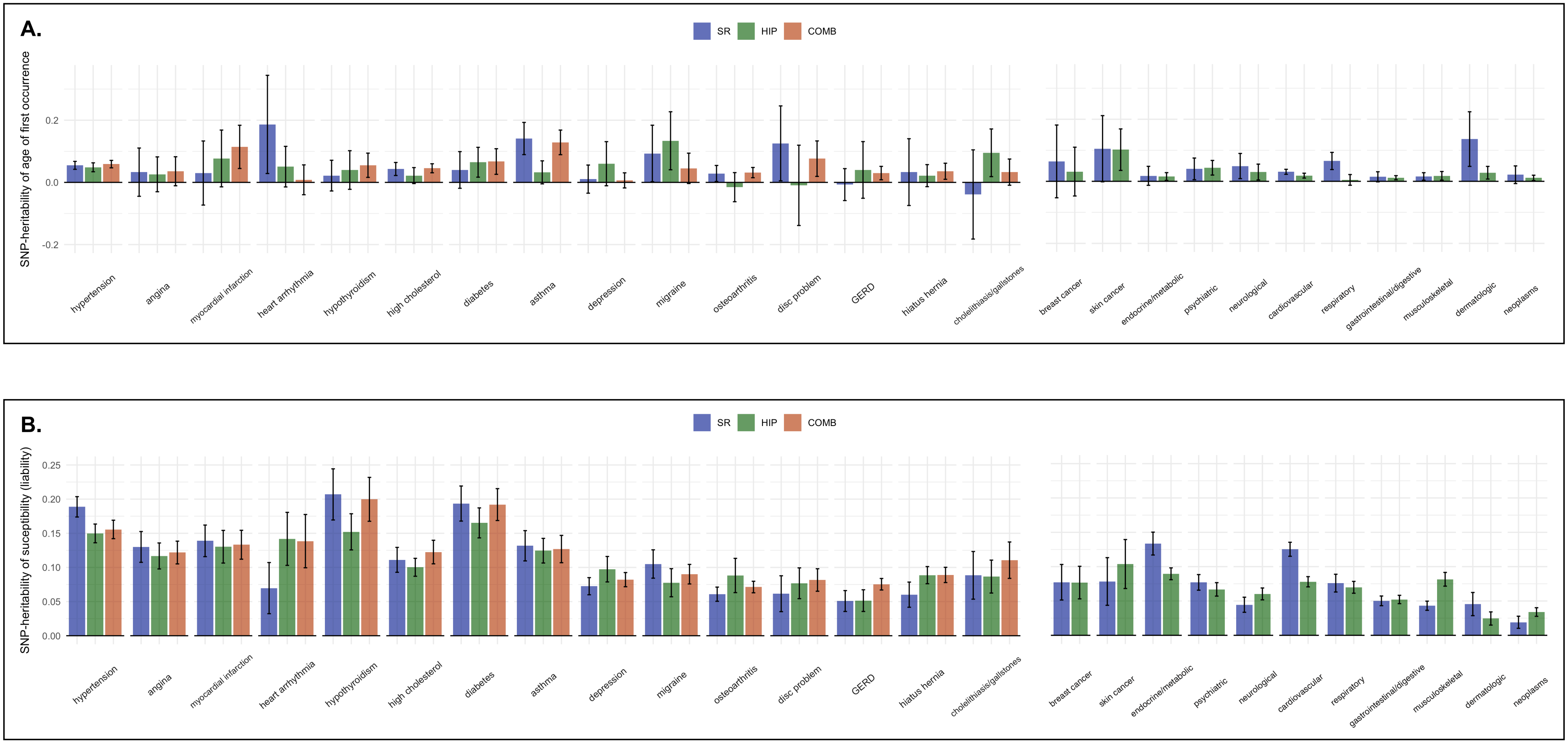


**Figure S10.** Comparison of heritability of age of first occurrence ($h^2_{aofo}$) and susceptibility ($h^2_{susc}$) for 26 traits with closely matched definitions between the analyzed UKBB datasets

Top panel depicts the $h^2_{aofo}$ estimates **(A)** along with the 95% confidence intervals (95% CI), while bottom panel shows the $h^2_{susc}$ estimates **(B)**. 15 of the 26 diseases phenotypes have a comparable definition across all three datasets (left), and an additional 11 phenotypes are matched only between the SR and the HIP datasets (right).

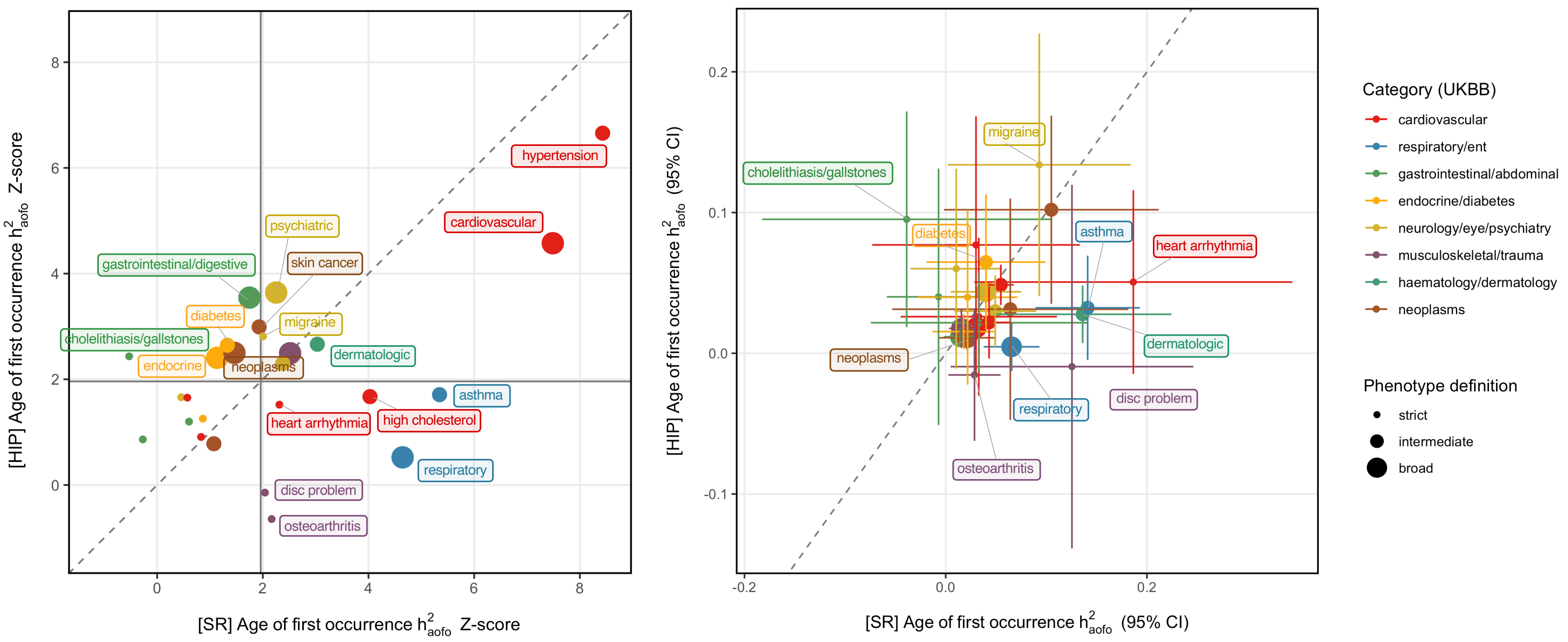


**Figure S11.** Comparison of $h^2_{aofo}$ for 26 mapped disease definitions in the SR and HIP datasets

Left panel shows the $h^2_{aofo}$ Z-scores in the SR (x-axis) versus in the HIP dataset (y-axis). Solid grey lines indicate Z-score = 1.96, above which the $h^2_{aofo}$ is significant at p-value < 0.05. Dotted grey line is the diagonal line for x = y.
Right panel shows the $h^2_{aofo}$ estimates with a 95% CI in each dataset. Labeled are traits with a significant $h^2_{aofo}$ at p-value < 0.05. Dotted line indicates x = y.
Each dot represents an individual disease, colored by disease categories in the SR dataset. The size of the dot corresponds how broadly defined the disease phenotype is.

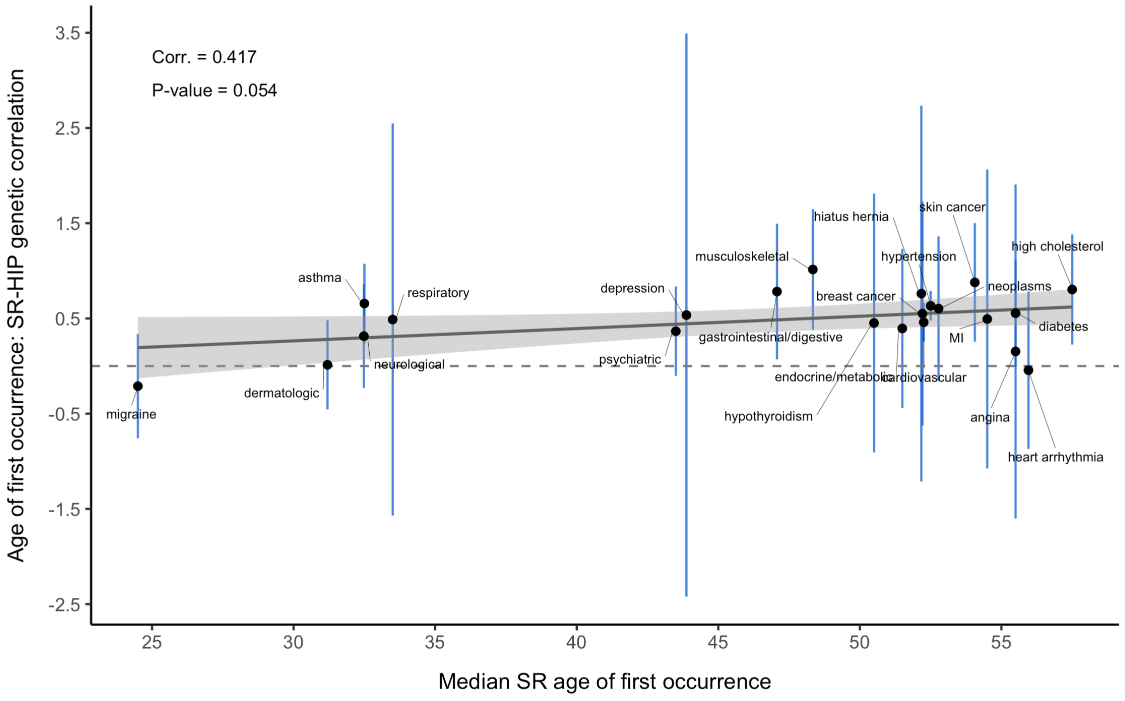


**Figure S12.** Genetic correlation ($r_g$) of age of first occurrence between SR and HIP endpoints increases slightly with median age of first occurrence across 26 mapped disease definitions

Median age of first occurrence in the SR dataset were plotted against the cross-dataset $r_g$ between SR and HIP for these 26 mapped diseases. $r_g$ was calculated using bivariate LDSR. Solid blue lines indicate the 95% CI for each $r_g$ estimate. The solid grey line denotes $r_g$ = 0.

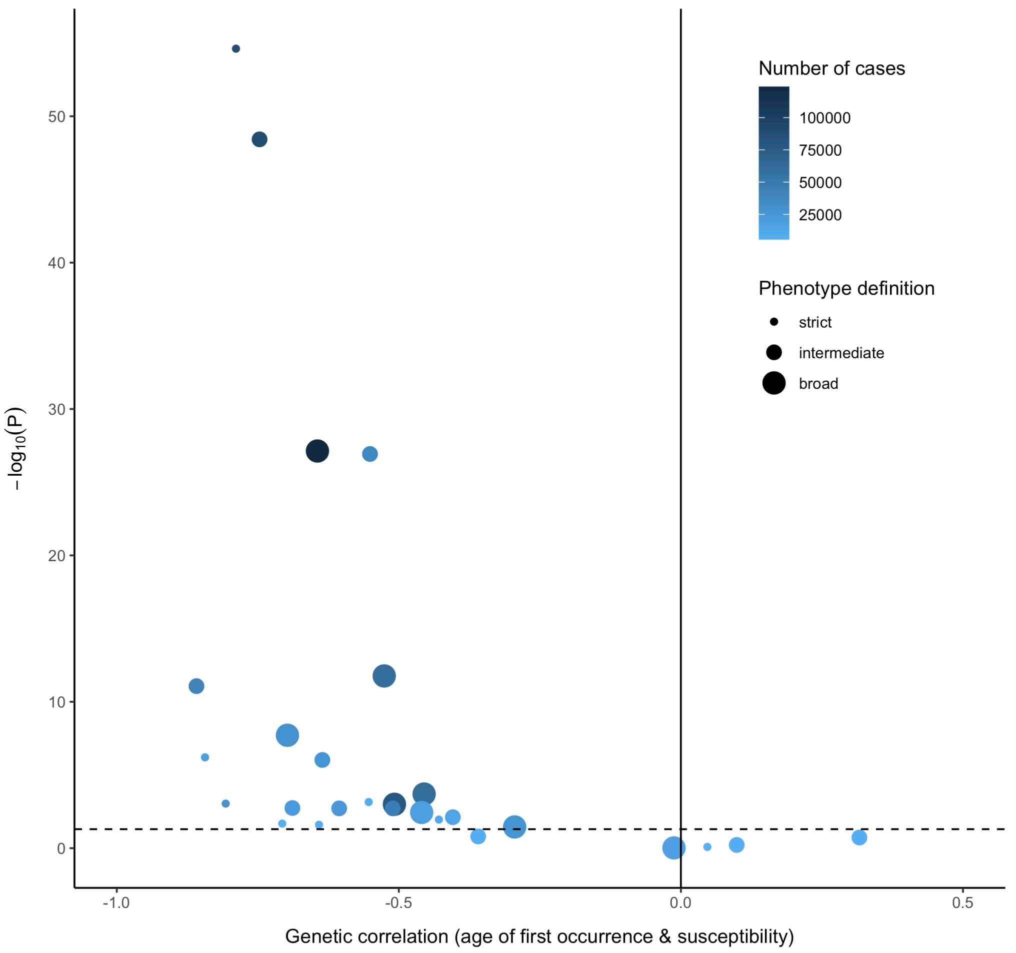


**Figure S13.** Genetic correlation ($r_g$) analysis is bound by power

Illustrated here are the results based on the SR dataset. $r_g$ was estimated using bivariate LDSR (x-axis) and plotted against p-values on the $-\log_{10}$ scale (y-axis). Each dot represents an individual disease, colored by number of cases in the SR dataset, with a darker color indicating a larger number of affected individuals. The size of the dot corresponds how broadly defined the disease phenotype is. The dotted line denotes the nominal significance threshold at $P$ = 0.05.

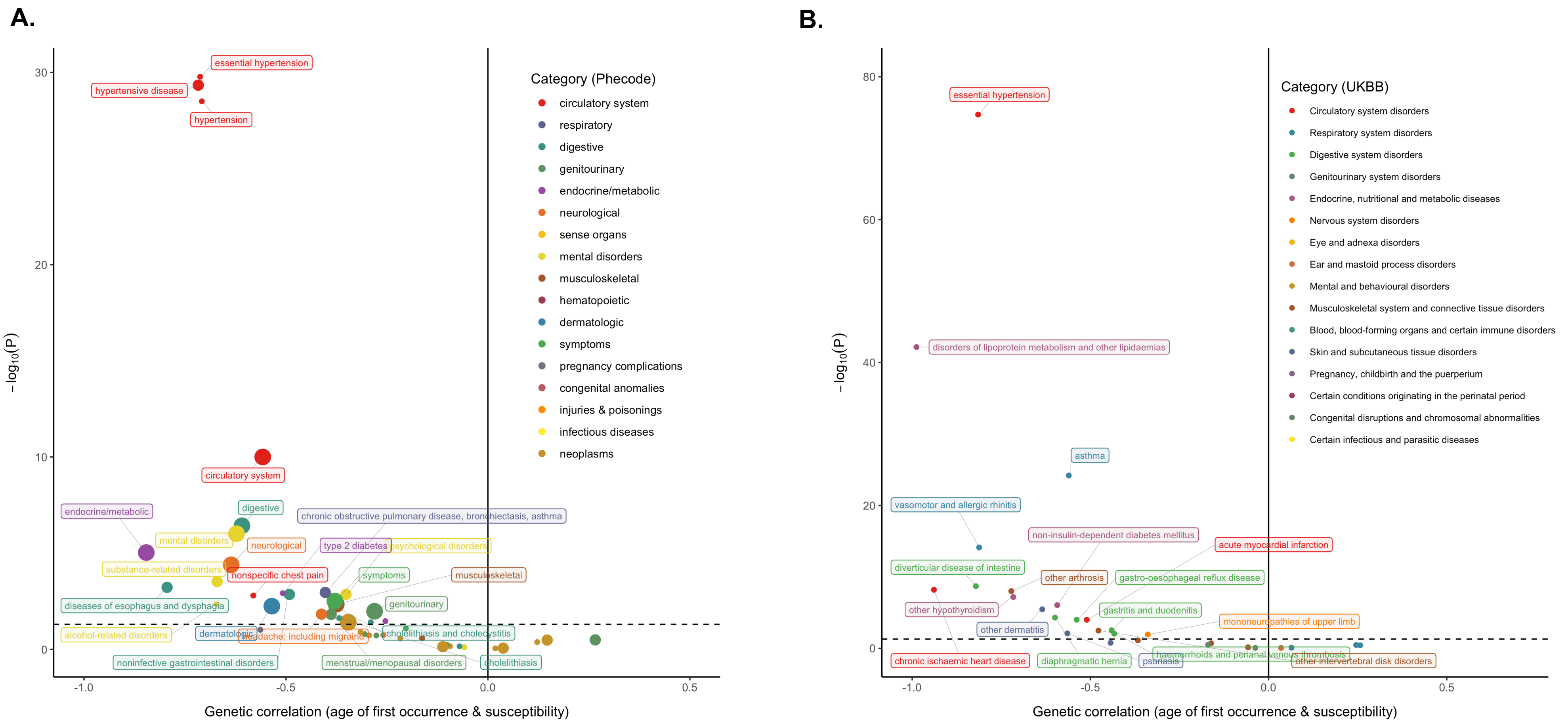


**Figure S14.** Genetic correlation ($r_g$) between age of first occurrence and susceptibility in the HIP dataset **(A)** and in the COMB dataset **(B)**

$r_g$ was estimated using bivariate LDSR (x-axis) and plotted against p-values on the $-\log_{10}$ scale (y-axis,). Each dot represents an individual disease, colored by disease categories used in the SR dataset; a larger dot corresponds to a broader disease definition. The dotted line denotes the nominal significance threshold at $P$ = 0.05. Labeled are conditions with a significant $r_g$ at FDR < 0.05. Analysis of SR diseases shows a similar pattern in Figure 2C.

**A.**
SR dataset

**B.**
HIP dataset

**C.**
COMB dataset

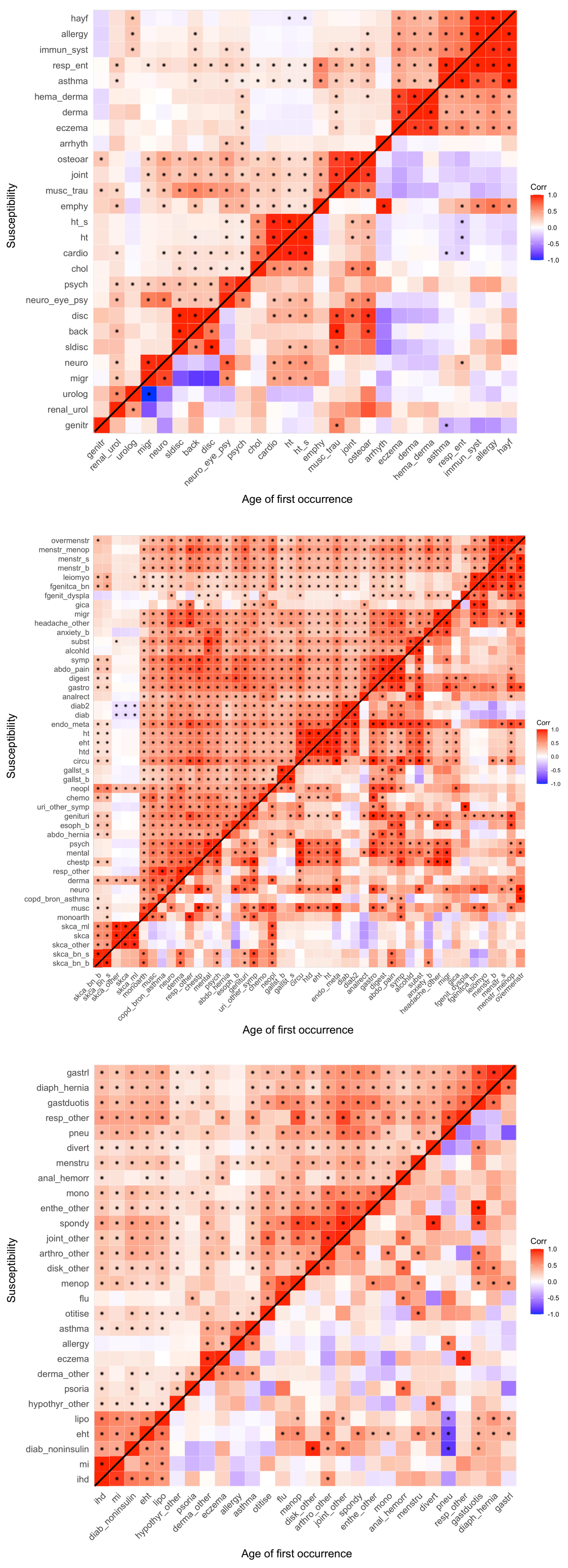


**Figure S15.** Genetic overlap between susceptibility endpoints and between age-of-first-occurrence endpoints in each dataset.

Pairwise genetic correlations ($r_g$) were estimated using bivariate LDSR for disease definitions that have a nominal significant $h^2_{aofo}$ and $h^2_{susc}$ . Upper triangle shows the hierarchical clustering of $r_g$ among the susceptibility endpoints, and the lower triangle shows the pairwise $r_g$ for age of first occurrence phenotypes given the same order in the upper triangle.

Significant $r_g$ is labeled with a star sign (“*”).

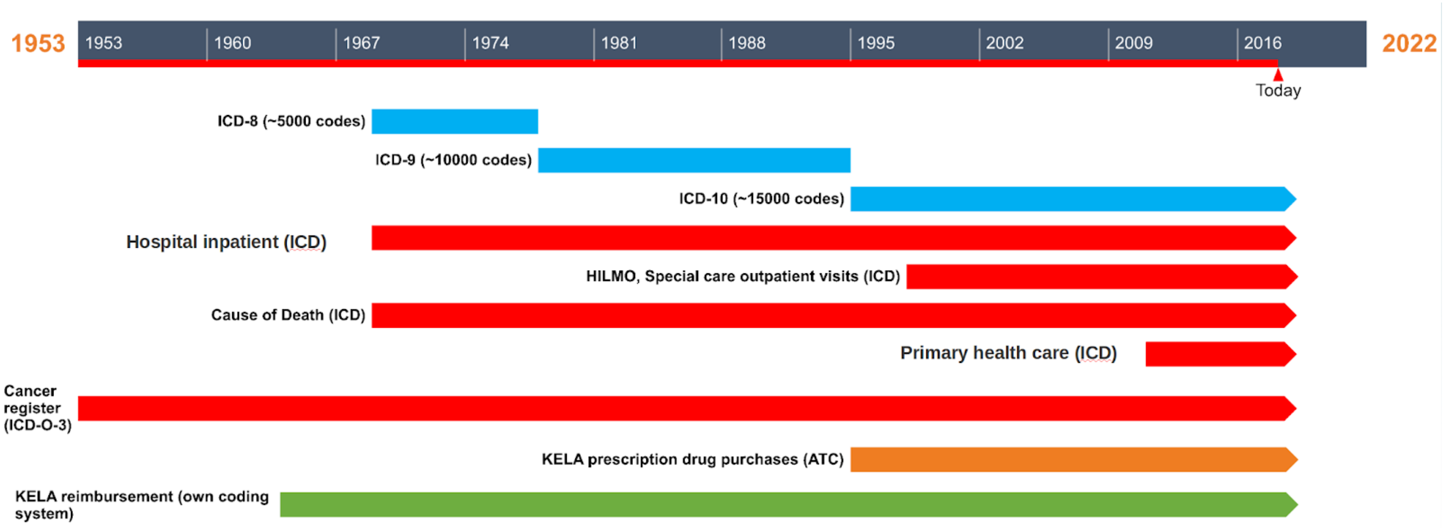


**Figure S16.** Time span covered by each ICD version and each Finnish nation-wide registry included in FinnGen

FinnGen disease endpoints are defined using nationwide registries, with data harmonized over the ICD revisions 8, 9 and 10, cancer-specific ICD-O-3, (NOMESCO) procedure codes, Finnish-specific Social Insurance Institute (KELA) drug reimbursement codes, and ATC-codes for medications. These registries span decades and are electronically linked to the cohort baseline data using the unique national personal identification numbers assigned to all Finnish citizens and residents.

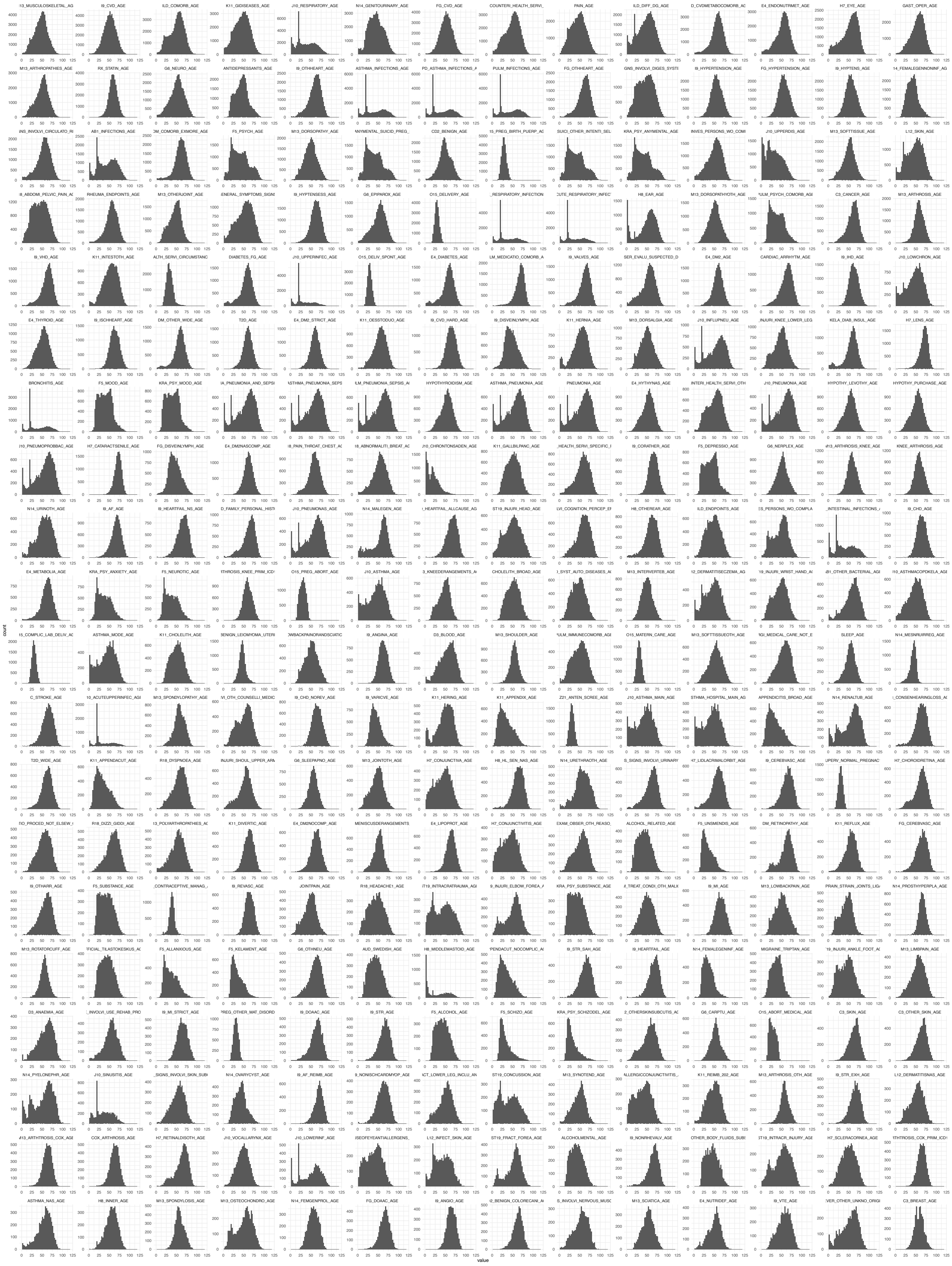

**Figure S17.** Age of first occurrence distribution for analyzed disease phenotypes in FinnGen

For each condition, age of first occurrence was defined as the earliest age of an event in the registries. Age of first occurrence varies across the analyzed diseases and ranges from 0 to 105.65 (median: 52.04, IQR: 28.22).

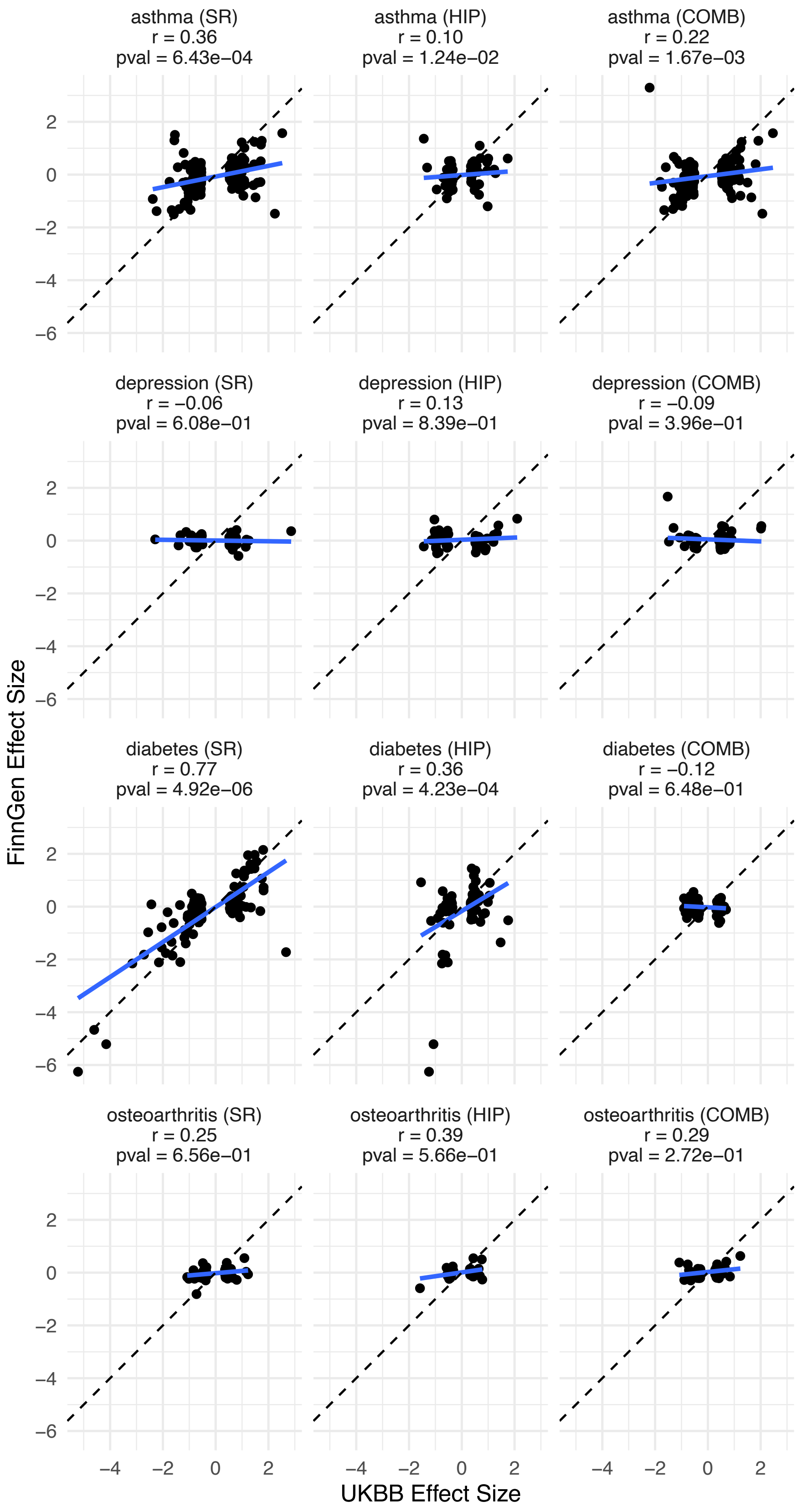


**Figure S18.** Effect size comparison in UKBB and FinnGen for selected disease phenotypes

Effect sizes (beta coefficient) for top loci ascertained in UKBB at a p-value < 0.0001 are compared against the effect sizes in FinnGen. The dashed line shows x = y, and the blue solid line shows the fitted regression line based on the data. A Pearson's correlation coefficient (r) was calculated along with a one-sample binomial (sign) test p-value (pval) to evaluate the concordance of the direction of the effects between the two cohorts.

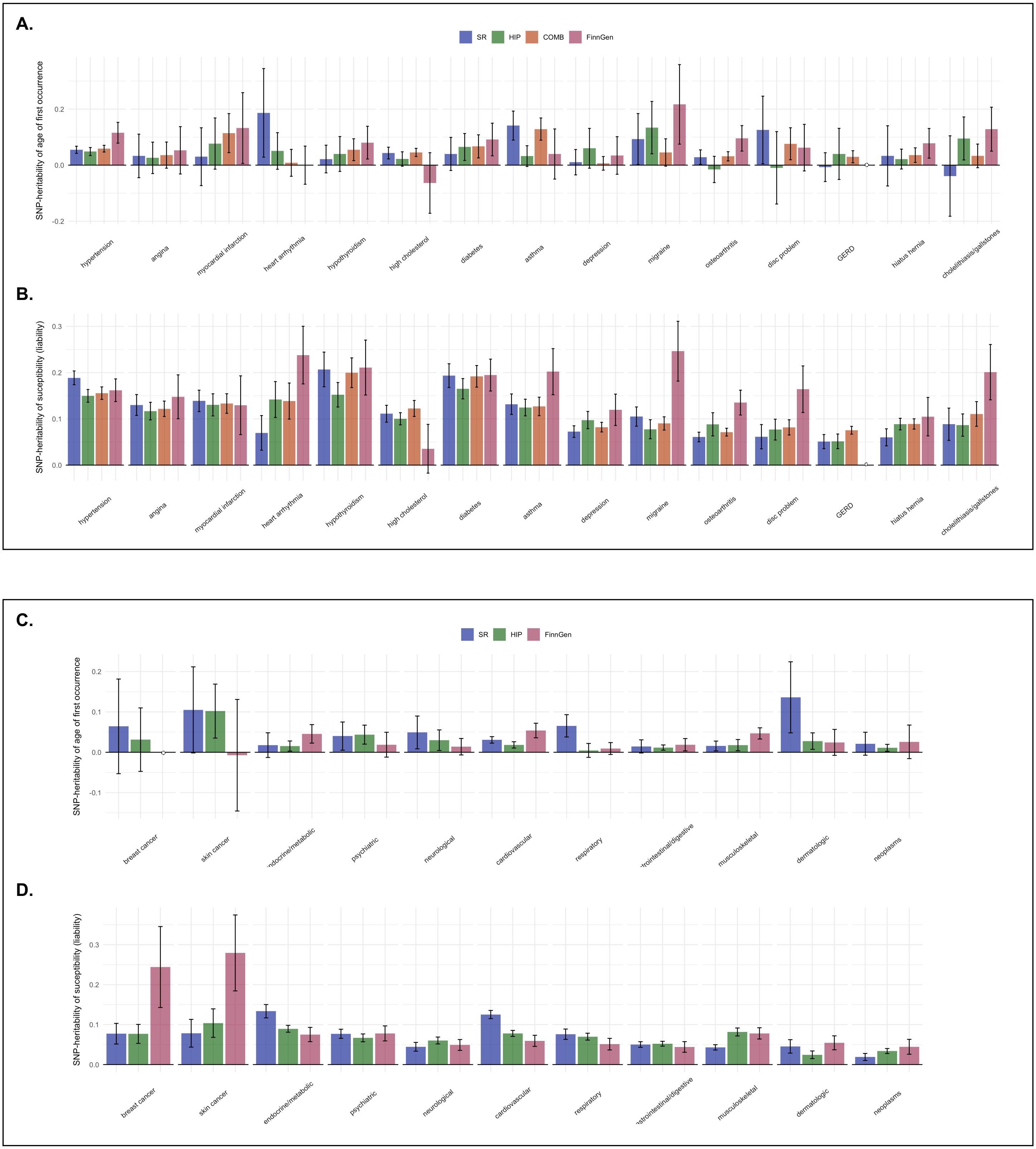


**Figure S19.** Comparison of heritability of age of first occurrence ($h^2_{aofo}$) and susceptibility ($h^2_{susc}$) and for 26 traits with closely matched definitions between the three UKBB datasets and FinnGen

Top panel depicts the $h^2_{aofo}$ **(A)** and $h^2_{susc}$ estimates **(B)** along with their 95% CIs for 15 diseases phenotypes matched across all datasets. Bottom panel **(C, D)** includes the estimates of an additional 11 phenotypes matched only between SR, HIP, and the FinnGen datasets.

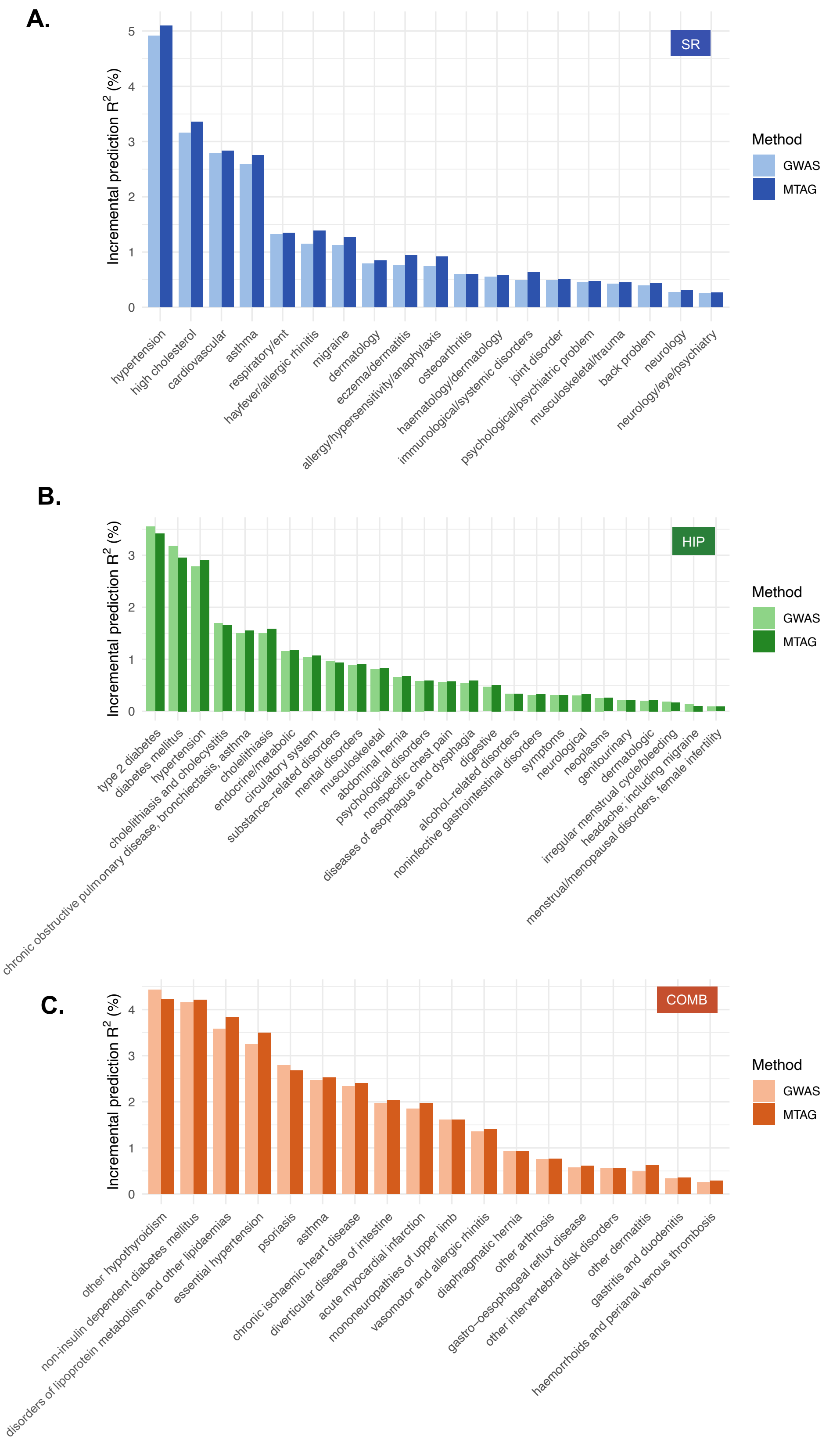


**Figure S20.** Polygenic risk prediction of MTAG versus GWAS in each UKBB dataset

For each dataset, an MTAG of age of first occurrence and susceptibility was performed for diseases with a significant negative $r_g$ between the two traits. PRS was then calculated using MTAG and GWAS results separately in a hold-out sample of 91K EUR individuals for diseases with >2000 cases. The incremental prediction $R^2$ (y-axis) for MTAG and GWAS was calculated as the increase in predictive power with PRS included over a covariate-only model.

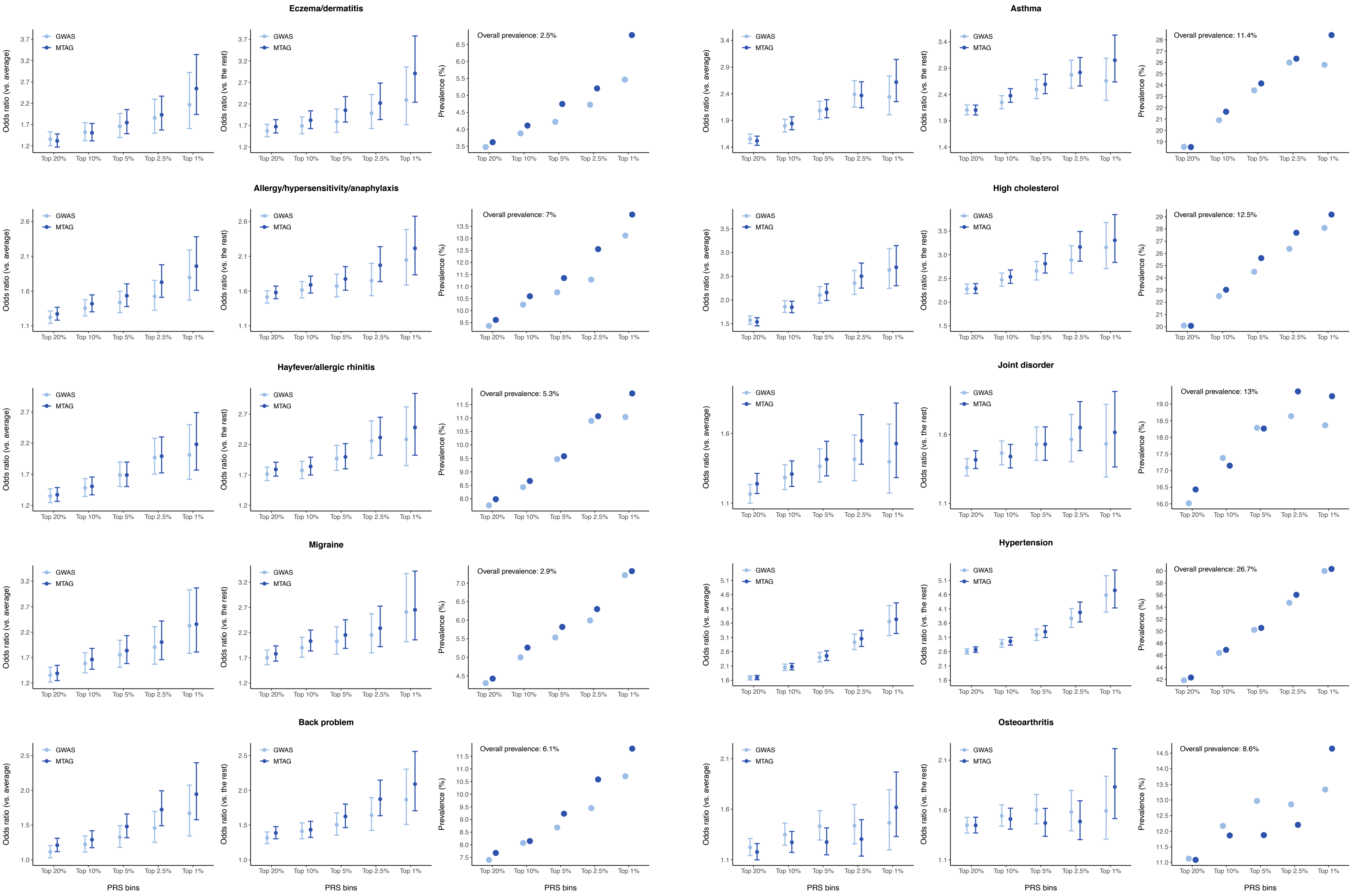


**Figure S21.** Performance of patient stratification using MTAG-PRS compared to GWAS-PRS for 10 individual diseases in the SR dataset
Left panel of each disease shows an adjusted odds ratio and its 95% CI for individuals who belonged to the top PRS percentile (1%, 2.5%, 5%, 10%, and 20%) versus those among the average percentiles (20-80%). Middle panel shows the adjusted odds ratio for top PRS percentiles relative to the rest of the sample. Right panel shows the proportion of cases identified in each of the top PRS percentiles computed using MTAG or GWAS summary statistics. Full results for all three datasets can be found in Tables S24-26.
