## Supplementary Notes for "Findings and insights from the genetic investigation of age of first reported occurrence for complex disorders in the UK Biobank and FinnGen"

### FinnGen cohort description

FinnGen is a public-private partnership project combining genotype data from Finnish biobanks and digital health record data from Finnish health registries (https://www.finngen.fi/en). Six regional and three country-wide Finnish biobanks participate in FinnGen. Finngen also includes data from previously established populations and disease-based cohorts.

*Phenotypes definition*

FinnGen disease endpoints are defined using nationwide registries. Data are harmonized over the International Classification of Diseases (ICD) revisions 8, 9 and 10, cancer-specific ICD-O-3, (NOMESCO) procedure codes, Finnish-specific Social Insurance Institute (KELA) drug reimbursement codes and ATC-codes for medications. These registries span decades, as shown in Figure S16, and are electronically linked to the cohort baseline data using the unique national personal identification numbers assigned to all Finnish citizens and residents.

*Genotyping*

Samples were genotyped with Illumina (Illumina Inc., San Diego, CA, USA) and Affymetrix arrays (Thermo Fisher Scientific, Santa Clara, CA, USA). Genotype calls were made with GenCall and zCall algorithms for Illumina and AxiomGT1 algorithm for Affymetrix data. Chip genotyping data produced with previous chip platforms and reference genome builds were lifted over to build version 38 (GRCh38/hg38) following the protocol described here: dx.doi.org/10.17504/protocols.io.nqtddwn. In sample-wise quality control, individuals with ambiguous sex, high genotype missingness (>5%), excess heterozygosity (+-4SD) and non-Finnish ancestry were removed. In variant-wise quality control, variants with high missingness (>2%), low HWE P-value (<1e-6) and minor allele count (MAC) <3 were removed. Chip genotyped samples were pre-phased with Eagle 2.3.5 (https://data.broadinstitute.org/alkesgroup/Eagle/) with the default parameters, except the number of conditioning haplotypes was set to 20,000.

*Genotype imputation with a population-specific reference panel*

High-coverage (25-30x) WGS data (N= 3,775) were generated at the Broad Institute and at the McDonnell Genome Institute at Washington University; and jointly processed at the Broad Institute. Variant callset was produced with GATK HaplotypeCaller algorithm by following the GATK best-practice for variant calling. Genotype-, sample- and variant-wise QC was applied in an iterative manner by using the Hail framework (https://github.com/hail-is/hail) v0.1 and the resulting high-quality WGS data for 3,775 individuals were phased with Eagle 2.3.5 as described above. Genotype imputation was carried out by using the population-specific SISu v3 imputation reference panel with Beagle 4.1 (version 08Jun17.d8b, https://faculty.washington.edu/browning/beagle/b4_1.html) as described in the following protocol: dx.doi.org/10.17504/protocols.io.nmndc5e. Post-imputation quality-control involved non-reference concordance analyses, checking expected conformity of the imputation INFO-values distribution, MAF differences between the target dataset and the imputation reference panel and checking chromosomal continuity of the imputed genotype calls.

*Ancestry assignment*

For principal components analysis, FinnGen data was combined with 1000 genomes data. Related individuals (<3rd degree) were removed using KING software. We considered common (MAF >= 0.05) high quality variants: not in chromosome X, imputation INFO>0.95, genotype imputation posterior probability>0.95 and missingness<0.01. LD-pruned ($r^2$<0.1) common variants were used for computing PCA with PLINK 1.92.
